# Forebrain speech networks specifically impaired in children with listening difficulties

**DOI:** 10.1101/2025.10.22.25338400

**Authors:** Hannah J. Stewart, Erin Cash, Lisa L. Hunter, Jonathan E. Peelle, Leanne Tamm, Stephen P. Becker, Jennifer Vannest, David R. Moore

**Affiliations:** Communication Sciences Research Centre, Cincinnati Children’s Hospital Medical Center, Cincinnati, Ohio, USA; Research in Patient Services, Cincinnati Children’s Hospital Medical Center, Cincinnati, Ohio, USA; Department of Psychology, Lancaster University, Lancaster, UK; Division of Psychology and Language Sciences, University College London, London, UK; Department of Otolaryngology-Head and Neck Surgery, University of Cincinnati, Ohio, USA; Department of Communication Sciences and Disorders and Department of Psychology, Northeastern University, Boston, Massachusetts, USA; Division of Behavioral Medicine and Clinical Psychology, Cincinnati Children’s Hospital Medical Center, Cincinnati, OH, USA; Department of Pediatrics, College of Medicine, University of Cincinnati, Cincinnati, Ohio, USA; Department of Communication Sciences and Disorders, University of Cincinnati, Ohio, USA; Division of Speech-Language Pathology, Cincinnati Children’s Hospital Medical Centre, Ohio, USA; Manchester Centre for Audiology and Deafness, University of Manchester, Manchester, M13 9PL, UK

**Keywords:** listening difficulties, pediatric auditory neuroscience, speech, sound, neurodevelopmental disorders

## Abstract

Children with neurodevelopmental disorders have a high rate of listening difficulty (LiD); impaired listening ability in the absence of hearing loss. However, the nature and mechanisms of LiD remain unclear. It has been proposed that LiD results from a primary auditory processing disorder, a perturbation of auditory specific brain activity underlying perception. An alternative hypothesis is that LiD results from a higher level, post-coding disorder of speech and language. This study used resting state fMRI of forebrain functional connectivity to distinguish between these hypotheses. It investigated the distribution of speech, non-speech ‘sound’, and visual processing networks in three groups of children aged 6-14 years with normal hearing. In addition to children with caregiver-identified LiD, two control groups were children diagnosed with attention-deficit/hyperactivity disorder (ADHD) and typically developing children. Relative to typically developing children, reduced functional connectivity of the speech network was found in children with LiD. No change was found in networks for non-speech sounds or visual stimuli in the children with LiD, suggesting a specific deficit in speech processing. Children with ADHD showed reduced connectivity in both speech and sound networks, but not in the visual network, suggesting a common underlying cause for auditory and speech difficulties. We conclude that listening difficulties in children are mediated by speech-specific neural mechanisms. The findings strengthen research calls for speech and language testing as a component of clinical pediatric audiological assessment.

**Significance statement:** This study provides compelling neurobiological evidence that childhood listening difficulties, without hearing loss, are linked to specific deficits in speech network connectivity rather than to a general auditory dysfunction or delayed development. Our findings highlight the value of caregiver observations. Questionnaire scores aligned more strongly than objective hearing measures with neurological differences, supporting caregiver perspectives in assessment. Persistent neural differences throughout childhood highlight the need for early identification and sustained intervention in listening difficulties. By bridging everyday observations with measurable brain function, this work has the potential to improve clinical approaches for neurodevelopmental disorders, informing more effective and evidence-based assessment and intervention strategies.

## Introduction

Around 10-20% of children and young adults and 30-50% of older people report hearing or listening difficulties (LiD) despite audiometrically normal hearing. These people typically struggle to perceive or separate elements of speech, especially in noisy environments (Mittelstadt et al., 2025; Moore et al., 2017; Moore, 2014; Moore, Hugdahl, et al., 2020; Motlagh-Zadeh et al., 2019; Su & Chan, 2017). The nature and mechanisms of LiD without hearing loss have long been a puzzle for speech, language and hearing clinicians, psychologists and researchers. A major problem has been codifying LiD, especially in young children. Led by research in developmental language disorders (Bishop, 2006; Bishop & McDonald, 2009) a caregiver-report instrument that has been rigorously validated in multi-site samples is now available to identify children with LiD (the ECLiPS; Barry & Moore, 2021; Denys et al., 2024).

Childhood LiD has high co-occurrence with both the symptoms and diagnoses of other neurodevelopmental disorders and hearing loss (Figure 1; Dillon & Cameron, 2021; Moore et al., 2018, 2025; Sharma et al., 2009; Tomlin et al., 2015). For example, compared to their typically developing peers, higher rates of LiD are seen in children with attention-deficit/hyperactivity disorder (ADHD) (Lanzetta-Valdo et al., 2017; Riccio et al., 1994) or developmental language disorder (DLD) (Ferguson et al., 2013, 2014). It has recently been found that more mild forms of hearing loss, not yet recognized clinically, are associated with LiD. These conditions are sometimes called “subclinical” (Mishra et al., 2022; Moore, Zobay, et al., 2020; Sharma et al., 2023) or “hidden” hearing loss (Liberman, 2015; Schaette & McAlpine, 2011). The failure of many neurodevelopmental disorder studies and clinics to test hearing adequately has also likely contributed to lack of reported associations (Moore et al., 2025).

**Figure 1:**
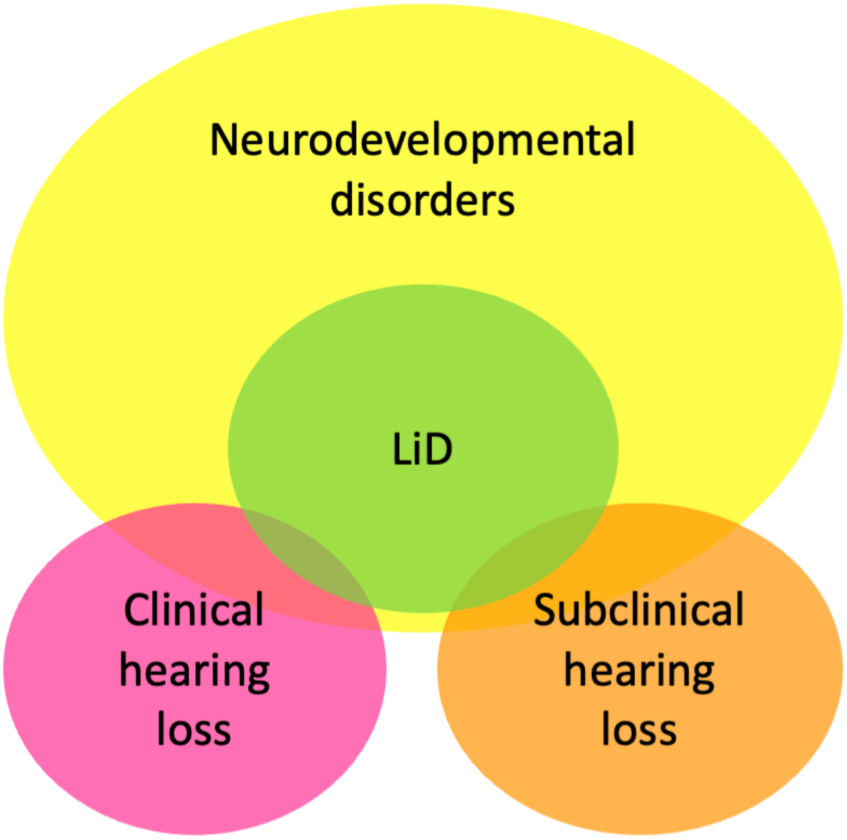
An illustration of different categories of hearing loss, listening difficulties and neurodevelopmental disorders. Children of all hearing and listening abilities can have comorbidity of neurodevelopmental disorders (e.g. ADHD, DLD, ASD and dyslexia). It is currently assumed that most children diagnosed with neurodevelopmental disorders are audiometrically normal (e.g., NIDCD, 2023). However, many neurodevelopmental disorder studies do not test hearing, and so the causality between neurodevelopmental disorders and hearing remains unclear. Children with LiD have high comorbidity with neurodevelopmental disorders (e.g. Moore & Hunter, 2013; Sharma et al., 2009; Tomlin et al., 2015. However, the causality between these two categories has neither been established or dismissed. Clinical hearing loss is defined as per the WHO guidelines of hearing thresholds below 20 dB; subclinical hearing loss is defined as hearing thresholds between 0 and 20 dB; and listening difficulties defined as hearing thresholds above 20 dB but with caregiver reports of everyday listening difficulties.

Previously, we recruited a group of 6-14 year old children with caregiver-reported LiD and a group of their typically developing (TD) peers. Both groups had normal hearing sensitivity (≤ 20dB HL, 0.25 - 16 kHz; Hunter et al., 2021). Nevertheless, the children with LiD had auditory perceptual deficits and wide-ranging neurodevelopmental disorders (Moore et al., 2018, 2020; Petley et al., 2021). These findings placed the source of LiD in either the central auditory nervous system, or in cortical structures that integrate auditory with other cognitive functions (e.g., attention, language, memory, vision) used for speech decoding and understanding.

Using resting state fMRI (Stewart et al., 2022) to examine the auditory cortex and temporal lobe of children with LiD, a close link was found between speech and language problems and reduced functional connectivity in children with LiD. Similar findings were reported by Alvand and colleagues (Alvand et al., 2022, 2023). However, the cortical markers of LiD may be more widespread across areas associated with neurodevelopmental disorders. In this study we examined whole-forebrain speech, non-speech “sound”, and visual processing networks in children with LiD or ADHD relative to TD children. We hypothesized that both LiD and ADHD groups would have altered connectivity of speech networks compared to their TD peers, supporting a common etiology for LiD across these neurodevelopmental disorders.

Using a meta-analytic approach, separate networks of regions of interest (ROIs) associated with processing “Speech” and “Vision” were identified from Neurosynth (Yarkoni et al., 2011). We independently developed a non-speech “Sound” network of ROIs for which a novel meta-analysis was generated. The sounds consisted of commonly used research auditory stimuli, such as tones (pure and complex), noises (white/pink/etc.), music and environmental sounds, to act as a control for the Speech ROI network. fMRI was used to examine resting state functional connectivity, assessing temporal correlations of fluctuations in the BOLD signal between ROIs and capturing interactions between brain regions in functionally associated networks (Biswal, 2012; Damoiseaux et al., 2006).

## Results

### Brain connectivity related to listening ability

Resting state functional MR images (rsfMRI) were obtained from the same MRI scanner for 39 TD children, 42 children with LiD, and 15 children diagnosed with ADHD (Figure 2A and B). All participants were audiometrically normal (Suppl. Figure 1), with the TD and LiD groups defined using a validated caregiver questionnaire, the Evaluation of Children’s Listening and Processing Skills (ECLiPS; Barry & Moore, 2021; Denys et al., 2024). Children with ADHD were primarily diagnosed using the caregiver-administered Kiddie Schedule for Affective Disorders and Schizophrenia (K-SADS; Kaufman et al., 1997).

**Figure 2:**
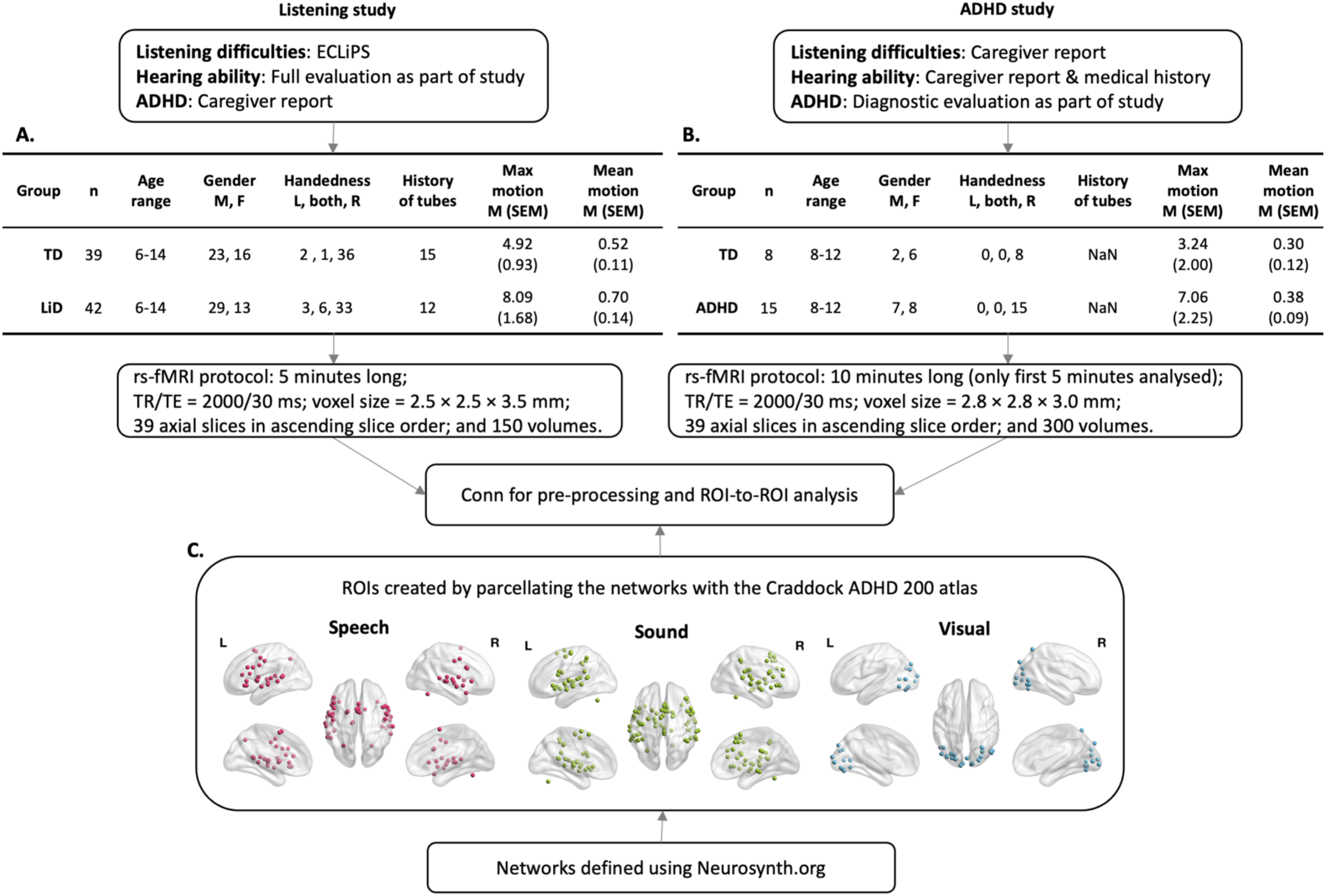
Methodology. In the listening study 81 children aged 6-14 years completed a large battery of behavioral and brain function tests. They all had “normal hearing” as defined by pure tone audiometry for the children with listening difficulty (LiD) and typical development (TD) (see Supp Figure 1 for audiograms). Children with LiD were recruited through clinical or family referral. TD and LiD were confirmed though a validated caregiver questionnaire (ECLiPS). Children with LiD performed poorly across a wide range of auditory (e.g. LiSN-S) and cognitive (NIH Toolbox) behavioral tasks (Petley et al., 2021, see also for full recruitment details). An additional group of children from the ADHD study were processed for comparison with the children tested for LiD. The children with ADHD had “normal hearing” as defined by parental report and medical records. We defined networks of regions of interest (ROI) across the cortex using the Neurosynth online resource with appropriate search terms for meta-analyses (see Methods for more). This created (C) 49 Speech, (D) 69 non-speech Sound and (E) 22 Visual ROIs, parcellated using the Craddock ADHD 200 atlas. Full ROI details are in Supp. Table 2. For visualization, BrainNet software was used to display foci in C (Xia, Wang & He, 2013). Max and mean motion were calculated with an outlier’s threshold of 0.9.

Speech, Sound and Vision networks of ROIs, defined using Neurosynth and parcellated with The Neuro Bureau ADHD-200 atlas (Bellec et al., 2017), are shown in Figure 2C. As expected, Speech and Sound networks were highly overlapping, but distinct across the superior temporal, frontal, and inferior parietal cortex. Visual networks, serving as a cortical control, were largely limited to the occipital cortex (Suppl. Figure 2).

#### Within network connectivity: Between groups

Among children with LiD, Speech network connectivity was notably sparse relative to TD children (Figure 3A). We used two approaches to quantify connectivity, both using FDR correction. First, a GLM controlling for age compared ROI-to-ROI pair connectivity between all ROI pairs within the Speech network (726 pairs at *p* < .05 as standard for all reported results; Suppl. Figure 5A). Second, ANOVA (F(2) = 73.35, *p* < .001, η^2^_p_ = .59; Table 1) and post-hoc Holm-corrected (family of 3) t-tests compared the averaged connectivity from across the whole Speech cortical network between groups (LiD vs TD t(79) = −11.94, *p* < .001, *d* = .35; Suppl. Figures 4, 6A; Suppl. Table 1). Both approaches showed a highly significant relative decline in connectivity in the LiD group.

**Figure 3:**
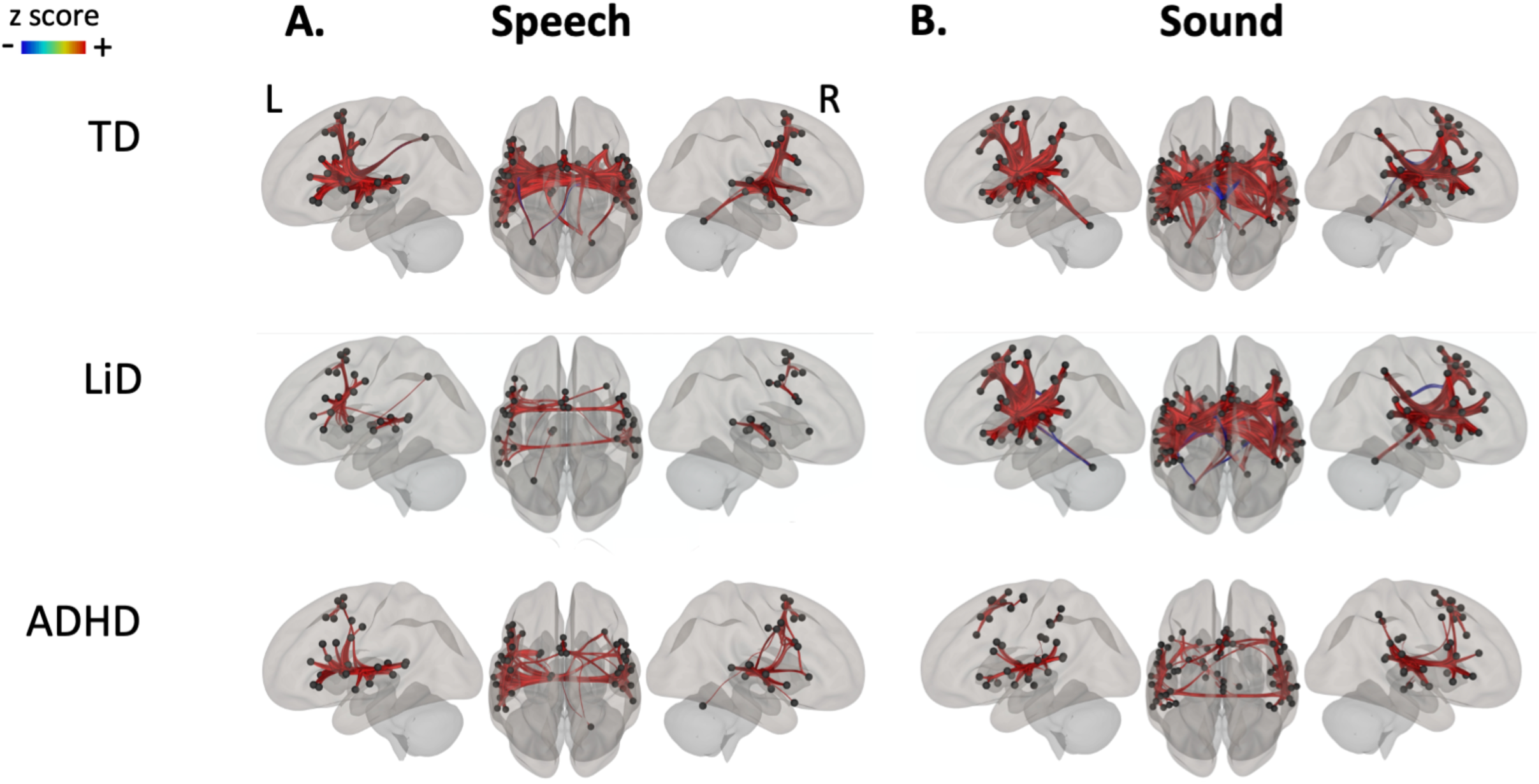
Highly specific differences in speech connectivity between children with and without listening difficulties were observed in direct quantitative comparisons between connectivity in the two groups. ROI-ROI resting state connectivity for each group of children in the (A) Speech, and (B) Sound networks. Thicker, more saturated color lines represent stronger connections between cortical areas. Anatomical representations of these ROI networks in full brain and brain slices are presented in Suppl. Figure 2. Heat maps further showing the quantitative strength of these connections can be found in Suppl. Figure 5. Large significant group differences were found in the (A) Speech network, especially between TD children and children with LiD. Minor group differences were found in the (B) Sound and Visual (Suppl. Figure 3) networks. Glass brain images are in neurological convention and were created in CONN functional connectivity toolbox (https://web.conn-toolbox.org).

**Table 1:** Significant group differences in averaged connectivity from across the Speech and Sound networks, but not the Visual network. Table shows group means and SD along with ANOVA results (FDR-corrected), see also Suppl. Figure 6 and Supp. Table 1 for post-hoc t-tests comparing between groups.

| Network | TD | LiD | ADHD | F(2) | p | $\eta_p^2$ |
| --- | --- | --- | --- | --- | --- | --- |
| Speech | .10 (.040) | .024 (.021) | .076 (.018) | 73.35 | < .001 | .59 |
| Sound | .028 (.013) | .028 (.013) | .021 (.005) | 3.68 | .029 | .068 |
| Visual | .011 (.004) | .010 (.003) | .011 (.003) | 1.16 | .32 | .023 |

Children with ADHD had an intermediate level of connectivity in the Speech network, with ROI-to-ROI pair analyses (Figure 3A; Suppl. Figure 5A) showing less connectivity than the TD group (136 ROI pairs at *p* < .05) but more than the LiD group (29 ROI pairs at *p* < .05). This pattern was also found for the ADHD group in the averaged connectivity across the Speech network (ADHD vs TD t(60) = −6.79, *p* < .001, *d* = .29; ADHD vs LiD t(63) = 3.40, *p* < .001, *d* = .46; Suppl. Figures 4, 6A; Suppl. Table 1).

The Sound network (F(2) = 3.68, *p* < .029, η^2^_p_ = .068, Table 1; Suppl. Figures 4, 6B; Suppl. Table 1) was, in contrast, almost indistinguishable between the LiD and TD groups. Connectivity between *all* Sound ROI-to-ROI pairs showed only two ROI pairs with significant group differences (*p* = .035 and *p* = .039; Figure. 3B; Suppl. Figure 5B). Averaged connectivity across the Sound cortical network found no LiD vs TD group difference (*p* = .81).

The ADHD group had a more sparsely connected and weaker Sound network compared to the other groups. These differences were significant as shown both by GLM assessing connectivity between all Sound ROI-to-ROI pairs (6 pairs for ADHD vs TD and 4 pairs for ADHD vs LiD, *p* < .05; Figure 3B; Suppl. Figure 5B), and averaged connectivity in the Sound network (ADHD vs TD t(60) = −2.53, *p* = .039, *d* = .28; ADHD vs LiD t(63) = 2.36, *p* = .040, *d* = .28; Suppl. Figures 4, 6B; Suppl. Table 1).

The Visual network (Figure 2C) had very similar connectivity between the three groups (Suppl. Figures 3, 6C), with just a single significantly different ROI-to-ROI pair when comparing the ADHD and LiD groups (*p* < .05). This was also reflected in the averaged connectivity analysis of the Visual network (*p* = .32; Table 1; Suppl. Figures 4, 6C).

To check for possible procedural differences between the listening and ADHD studies, we compared network connectivity between age-matched TD children in each study using a GLM (Suppl. Figure 7A). Minor significant connectivity differences were found in six ROI pairs in the Speech network (*p* < .05); and six ROI pairs in the Sound network (*p* < .05). No differences were found in the Visual network.

We also compared network connectivity between children with LiD, but with (n = 11) or without (n = 31) a diagnosis of auditory processing disorder (APD; Suppl. Figure 7B). APD is a widely used diagnosis, especially in pediatric audiology, and a hypothesized source of LiD (Dillon & Cameron, 2021; Petley et al., 2021). No significant connectivity differences were found in the Speech or the Visual network, but two ROI pairs in the Sound network were significantly stronger in LiD children with an APD diagnosis (p < .001).

Finally, we compared children with LiD and with (n =11) or without (n = 31) referrals to speech, language pathology (Suppl. Figure 7C). Results showed 15 ROI pairs with group differences in the Speech network (*p* < .05), two ROI pairs in the Sound network (*p* = .048) and none in the Visual network.

### Processing along the auditory cortical pathway

To examine effects of LiD within classically-defined central auditory system components of the Speech network, we measured connectivity between ROI pairs along the auditory forebrain pathway for the TD and LiD groups. Pairs were first examined between the left thalamus and bilateral primary auditory areas (Heschl’s gyrus). Results showed stronger connectivity between primary auditory areas (left thalamus and left Heschl’s gyrus; Figure 4A) for the TD group compared to the LiD. The right thalamus was not highlighted as a ROI in Neurosynth and so was not included in this analysis. As connections between the medial geniculate body (principal nucleus of the auditory thalamus) and primary auditory cortex are homolateral (Malmierca & Hackett, 2010) it is possible that a similar pattern would be found between the right thalamus and right Heschl’s gyrus.

**Figure 4:**
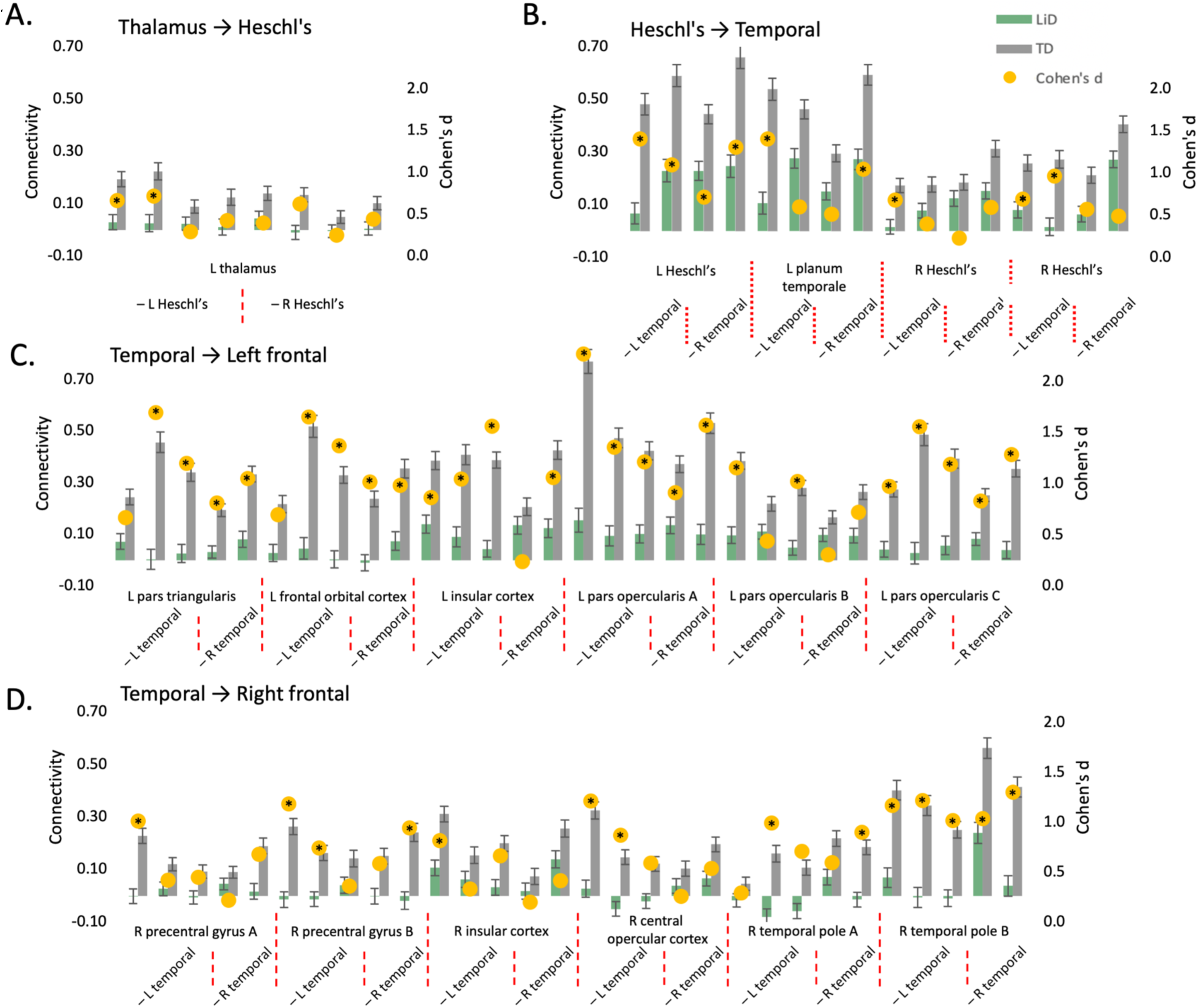
There was a higher proportion of TD dominance in connectivity strength between ROI pairs within the Speech network as we moved from primary auditory cortex to secondary auditory cortex to frontal-temporal areas. Strength of ROI-to-ROI connectivity (left axis) per group (LiD green, TD grey) and effect sizes (yellow dots, right axis) of the group comparisons corrected for age between: (A) basic auditory processing areas: left thalamus (ROIs 67, 113) to left (ROIs 144, 176) and right (ROIs 7, 185) Heschl’s gyrus; (B) secondary auditory cortex areas: left (ROIs 144, 176) and right (ROIs 7, 185) Heschl’s gyrus to left (ROIs 109, 197) and right (ROIs 22, 70) temporal areas; and frontal-to-temporal areas, moving in a dorsal-posterior direction, between left (ROIs 144, 197, 109) and right (ROIs 22, 70) temporal areas to (C) left frontal (ROIs 168, 48, 124, 157, 66, 179) and (D) right frontal (ROIs 60, 94, 59, 129, 110, 155) areas. Significant group differences in connectivity strength were found in: (A) two out of eight ROI pairs, both within the left hemisphere; (B) 9 out of 16 ROI pairs; (C) 24 out of 30 ROI pairs; and (D) 14 out of 30 ROI pairs. * indicates ROI pairs with a significant group difference, *p*-values were respectively adjusted for the following number of comparisons: 8, 16, 30 and 30.

We next examined connectivity of ROI pairs between the primary and secondary auditory cortical areas bilaterally (Heschl’s gyrus and planum temporale in the posterior, superior temporal gyrus). Here, we found a higher proportion of pairs where the TD group had stronger connectivity (Figure 4B) compared to the thalamus and primary AC pairs. Primary-secondary AC pairs that showed significant group differences mainly involved the left hemisphere, with left Heschl’s and planum temporale connecting to the left supramarginal gyrus and bilateral superior temporal gyrus. Significant connections from right Heschl’s were with left supramarginal gyrus and left superior temporal gyrus.

Finally, we examined ROI pairs between bilateral secondary auditory temporal areas and bilateral temporo-frontal cortical areas used for phonological processing, language recognition, language production, working memory, and social/emotional understanding (bilateral temporal gyrus; left pars triangularis, frontal orbital cortex, and pars opercularis; and right temporal pole, central opercular cortex, insular cortex and precentral gyrus). The left frontal connections showed the strongest group differences in connectivity strength (Figure 4C). TD dominance was found in almost all of the pairs (83% from the left temporal and 75% from the right temporal), with non-significant group differences in connections between left temporal areas and left pars triangularis, frontal orbital cortex and pars opercularis, and right temporal areas and left insular cortex and pars opercularis. Comparatively, the right frontal connections only showed a TD dominance in 56% of connections with the left temporal and 25% with the right temporal areas (Figure 4D). Significant group differences in connections were mainly between bilateral temporal areas and right temporal pole, and between left temporal and right frontal areas (precentral gyrus, insular cortex and central opercular cortex).

### Brain connectivity related to hearing and cognitive skills

In the larger listening study for which these LiD and TD groups were recruited, extensive behavioral data were available (Hunter et al., 2021, 2023; Moore et al., 2025; Moore, Hugdahl, et al., 2020; Petley et al., 2021, 2024). Brain-behavior relationships were analyzed for the ROI pairs from the Speech network that had shown significant group differences in the previous auditory pathway analysis (* ROI pairs in Figure 4; Supp. Table 4).

Significant positive correlations were found between connectivity strength and listening (both subjective - ECLiPS, and objective - SCAN), and between connectivity strength and cognitive skills (NIH) across the TD children and children with LiD. However, connectivity strength and speech-in-noise skills (LiSN-S) were not significantly correlated (Figure 5, Supp. Figure 8). Both the strength and significance of brain-behavior correlations increased as the ROI-to-ROI pairs moved rostrally. Connectivity z-scores from ROI pairs between the left thalamus and primary auditory cortical areas showed positive correlations with listening skills (subjective and objective) but not for speech-in-noise and cognition (.2 ≤ *r* ≤ .40, *p* ≤ .027; Figure 5A; Supp. Figure 8A). Connectivity z-scores from ROI pairs between primary and secondary auditory cortical areas were significantly correlated with listening skills (consistently for the ECLiPS, and sporadically for the SCAN) and cognition, and more uniformly so for pairs including the left primary auditory cortex (.22 ≤ *r* ≤ .5, *p* ≤ .049; Figure 5B; Supp. Figure 8B). Temporo-frontal cortical connectivity z-scores showed very different patterns of results for left and right frontal areas. The connections between bilateral temporal and left frontal areas showed strong positive correlations with listening skills (subjective and objective) and cognition (.23 ≤ *r* ≤ .58, *p* ≤ .047; Figure 5C; Supp. Figure 8C). However, connections between bilateral temporal and right frontal areas showed weaker correlations, and only with subjectively measured listening skills (ECLiPS) and, sporadically, with cognition (.22 ≤ *r* ≤ .51, *p* ≤ .048; Figure 5D; Supp. Figure 8D). These right frontal area connections did not show any correlations with objectively measured listening skills (SCAN).

**Figure 5:**
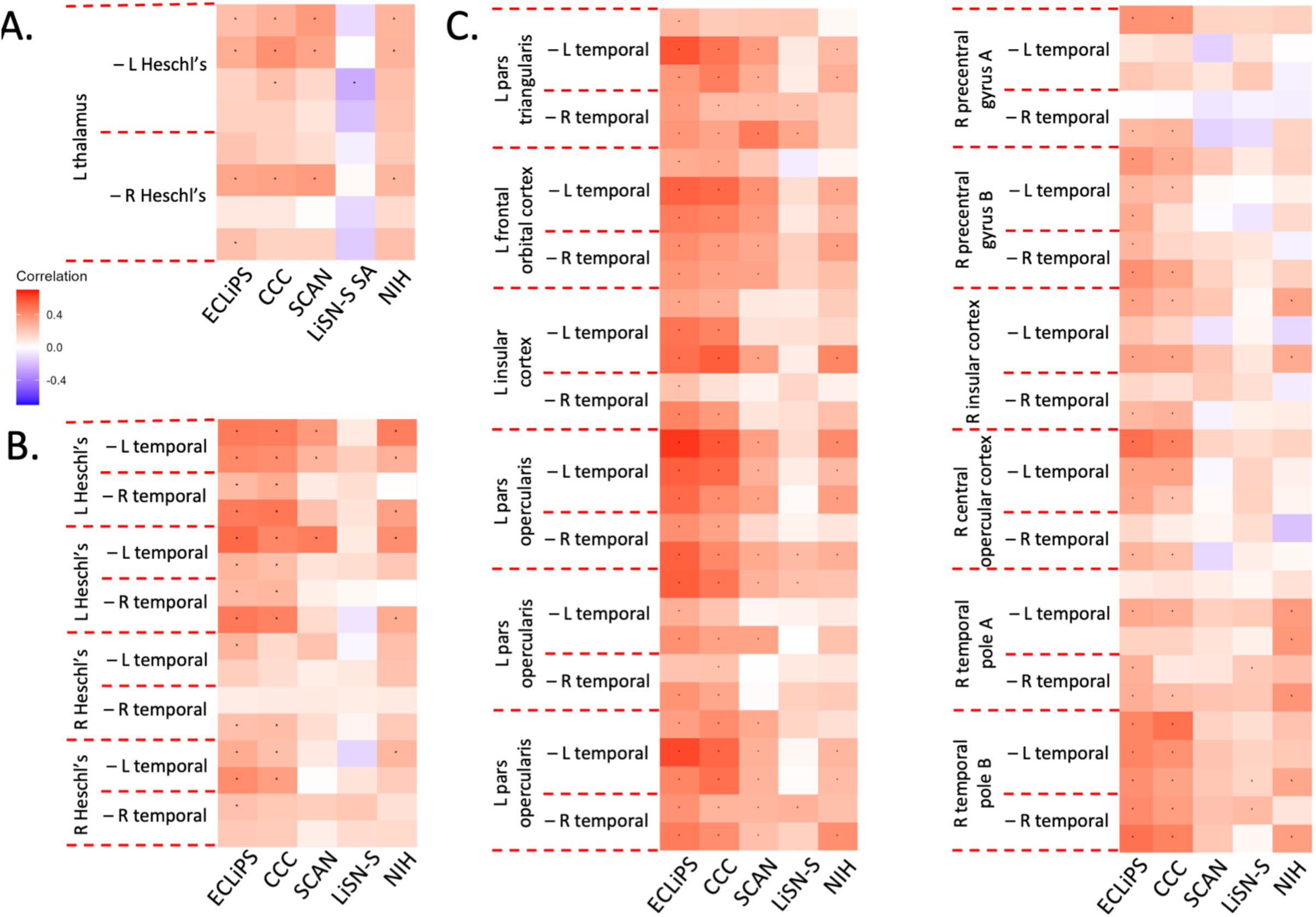
Significant brain-behavior correlations in the Speech network increased as we moved from primary auditory cortical areas to secondary auditory cortex areas to frontal-temporal areas. Brain-behaviour correlations were conducted on ROI-to-ROI pairs that showed significant group differences between TD and LiD groups (see Fig 4). Significant positive brain-behaviour correlations were found between (A) primary auditory cortical connectivity and general listening skills, but not for speech in noise and cognition; (B) secondary auditory cortical connectivity and listening skills and cognition; (C) temporo-left frontal cortical connectivity and listening skills (questionnaire and behavioural) and cognition, but not for speech-in-noise; and (D) temporo-right frontal cortical connectivity and listening skills (questionnaire) and cognition, but not for listening skills (behavioural) and speech-in-noise. The scatters detailing individual data points are in Supplementary Figure 8.

### Effect of age on brain connectivity

Each participant group had different patterns of changing connectivity with age in the Speech and Sound networks, and only the TD children showed any age-related changes in the Visual network (Suppl. Figure 9). These observations were confirmed by a GLM with FDR-correction (all groups *p* < .05) comparing ROI-to-ROI connectivity between all ROI pairs and ages separately for each group (TD and LiD: 6/7, 8/9, 10/11, 12/13/14 y.o.; and ADHD 8/9, and 10/12 y.o.). Planned post-hoc GLMs comparing age ranges (Supp. Table 5) in the TD group highlighted age-related (6/7 to 8/9 y.o.) right hemisphere dominated changes (STG, planum temporale, precentral gyrus, IFG). In the Sound network, TD children between 6/7 to 10/11 y.o. showed decreases in connectivity between the left putamen and right limbic areas (cingulate gyrus), and within the right limbic regions. In the older age ranges, there were similarities in the development of the Speech and Sound networks for the TD children, with increases in connectivity between and within bilateral primary and secondary auditory cortical areas. After age 10/11 y.o. there were no further changes in connectivity in the Speech network, but the Sound network continued to increase.

The LiD group showed contrasting and slower development in both Speech and Sound networks. Strikingly, connectivity strength in this group *decreased* significantly after 10/11 y.o., as seen in the Speech network between auditory and fronto-temporal areas involved in phonological processing and semantic judgements (left pars triangularis and pars opercularis). In the Sound network, between 8/9 to 10/11 y.o., connectivity increased between subcortical left pallidum and right limbic areas (paracingulate gyrus), within limbic areas (right paracingulate gyrus and left insular cortex) and between left secondary auditory cortex and right middle frontal gyrus. As in the Speech network, children with LiD went on to show a decrease in Sound network connectivity, but later, at 12/13 y.o., and between bilateral primary auditory cortex, right limbic regions and parietal areas. Again contrasting with TD children, children with LiD showed no more changes in connectivity in the Sound network after 10/11 y.o.

The ADHD group showed yet another growth pattern in the Speech network, with no significant age related changes *until* ages 10/12 y.o. and between the secondary auditory and fronto-temporal cortical areas. In the Sound network there were very minor increases in connectivity between 8/9 to 10/12 y.o.

## Discussion

In this study we examined whole forebrain functional connectivity of temporal-correlations within three networks (Speech, Sound, and Visual) defined by meta-analysis. Two clinical groups (LiD and ADHD) had altered temporal connectivity compared to TD children, but also to each other and across age.

The study took a different approach than previous cortical MRI studies investigating listening by using resting state fMRI and a data-driven methodology to delineate Speech and Sound networks, alongside a control sensory network - Vision. The meta-analysis used activation likelihood estimation (Laird et al., 2011; Yarkoni et al., 2011) - a powerful methodology not limited to a specific task paradigm (e.g., speech recognition or oddball). Instead, the identified networks represented the cortical areas used across the whole brain to process auditory (speech and sound) and visual stimuli. The connectivity analysis thus allowed assessment of what areas were working in sync (high connectivity), and whether there was a decoupling between areas (low connectivity).

### Children with listening difficulties

Children with LiD showed a dramatic and wide-ranging reduction in Speech cortical connectivity compared to TD children and those with ADHD. These data strongly suggest that children with LiD have a speech-listening cortical deficit. Relative to TD children, children with LiD showed very minor differences in their Sound cortical network and none in their Visual cortical network. This clearly highlights a specific, cortical speech-listening deficit for children with LiD, rather than a more general auditory processing deficit. This interpretation ties in with previous behavioral results showing that these children with LiD, despite normal audiometry (Hunter et al., 2021), had impaired performance on auditory tasks involving speech (Hoyda et al., 2025; Kojima et al., 2024; Petley et al., 2021).

At the subcortical level, there is previous evidence for impaired temporal processing of speech signals in children with LiD (Kraus & Anderson, 2016) and altered auditory evoked responses in children with neurodevelopmental disorders (Goswami, 2022). We previously reported that the children with LiD in the current study had significantly increased acoustic reflex growth and slightly reduced auditory brainstem evoked electrical response latencies that correlated with speech in noise and behavioral temporal processing problems (Hunter et al., 2023; Petley et al., 2024). Together, these studies suggest subtle functional changes in brainstem and cortical processing in children with LiD. Combined with the increasing strength of brain-behaviour relationships in the Speech network (discussed below) from the thalamus to the temporal cortex, and then to the frontal areas, these data suggest that children with LiD have subcortical auditory processing similar to TD children.

The data add to the more punctate differences found at the cortical level in recent MRI studies of children with listening difficulties compared to their typical listening peers. While the same brain areas were identified in groups of children with and without LiD during speech recognition task-based MRI, there were clear group differences in temporal connectivity between these brain areas (analysed on separate rsMRI data due to analysis restrictions from the SPARSE/Hush paradigm used to reduce the effects of scanner-related noise, Hall et al., 1999; Vannest et al., 2009). Group differences were specifically between ROIs identified for the later stages of speech listening (intelligibility and semantics), not between ROIs identified for earlier speech listening (phonology) (Stewart et al., 2022). Furthermore, multimodal regional differences have been identified at the group level (Alvand et al., 2022, 2023). Along with this study, these patterns of results are markedly different from the global structural and functional changes observed in children with profound deafness (Cui et al., 2022; Huang et al., 2015; Li et al., 2019; Park et al., 2018; Shi et al., 2016).

### Children with ADHD

Children with ADHD showed decreased connectivity in both the Speech and Sound networks compared to TD children. However, their Speech cortical connectivity was stronger than children with LiD. We propose that the similarities in maturation of both the auditory networks within the ADHD group may reflect changes in bottom-up processes. Unfortunately, detailed audiometric analysis of these children was not available; a common shortcoming of many neurodevelopmental assessments. Although the age range was limited for the ADHD group, there was a small increase in connectivity of both Speech and Sound networks between age groups, meaning similar under-development and growth in both of these auditory networks.

Children with ADHD often exhibit poorer-than-typical speech and language skills, proposed to be impacted by delayed development of cognitive skills, such as executive function (Hawkins et al., 2016). Therefore, the weakness in the Speech network was anticipated, likely reflecting a life-long difficulty attending to disconnected words and conversations. However, the weakness in the Sound network was unexpected, since reactions to non-speech sounds might be expected to be more subcortical and not subject to the same non-sensory cortical influences that influence speech processing.

It is possible that weaker connectivity in multiple cortical networks contributes to poorer attention. However, reduced functional connectivity was not seen for the ADHD group in the Visual network. Weaker connectivity in the Sound network may instead relate to reported benefits in focus when inattentive children listen to task-irrelevant white noise while completing executive function tasks (Helps et al., 2014). Current theories discussing why task-irrelevant noise (usually non-speech sounds) can help children with ADHD propose that bottom-up processes may contribute to problems attending and listening by these children (see Pickens et al., 2019 for review).

The distinctiveness between the ADHD and LiD groups appears to be influenced by differences in their cortical language and cognitive machinery. Neurodevelopmental disorders, including LiD (Moore & Hunter, 2013), have often been suggested to have more difficulties in common than they do separately. However, the results presented here are more consistent with a transdiagnostic approach (Astle et al., 2022). According to this approach, while children with LiD and ADHD may both present with difficulty listening, their listening is underpinned by different auditory neural networks. The current data suggest that children with ADHD may have generally under-developed cortical processing of auditory information.

### Typically developing children

TD children had rich functional connectivity through all three cortical networks. Several connectivity properties could be predicted from adult literature (e.g. Speech: Hickok, 2012; Sound: Lewis et al., 2011; Visual: Bullier, 2001) and are consistent with previous regional activation data (Stewart et al., 2022). In the Speech and Sound networks, connectivity was distributed across the left and right temporal, frontal and parietal cortex, and through the corpus callosum, with relatively minor but nevertheless distinctive changes during development from age 6-13. The Visual cortical network was also richly connected by age 6 and showed very minor changes with age.

Age-related connectivity changes in the Speech network were focused in the right hemisphere between ages 6/7 to 8/9, including the primary auditory cortex, motor cortex and inferior frontal gyrus (associated with a wider inhibition network, Hampshire et al., 2010). Development of connectivity between the left auditory areas and motor cortex continued through to 12/13/14 y.o., consistent with the development of speech lateralization during childhood through either language learning or maturation (Holland et al., 2007; Kadis et al., 2011; Karunanayaka et al., 2007; Sroka et al., 2015; see Enge et al., 2020 for a meta-analysis). Together, these observations provide a reality check for the current data, both across and within ages.

Compared to speech processing, the organization and development of sound processing networks is much less understood. Data from the current study’s TD children extend our understanding of the development of parietal operculum (PO) connectivity to sound areas, showing stronger connectivity with age between the left PO and right supramarginal gyrus, important for rhythm memory (Oberhuber et al., 2016; Schaal et al., 2017). The PO is the most distributed sensory processing neurocircuit (Jung et al., 2009) with bilateral connections across all five lobes. Direct electrical stimulation in adults has demonstrated clear asymmetries, the left frontal and parietal PO showing associations with language and motor, respectively, and the right temporal PO with auditory function (Mălîia et al., 2018). Previous developmental work has shown that functional connectivity between language areas (left IFG pars triangularis) and right PO are stronger in 16-18 year old typically developing adolescents compared to 4-6 year old children (Youssofzadeh et al., 2018). However, both the TD children and children with LiD showed the opposite specialisations for the left and right PO compared to adult results (Mălîia et al., 2018; Youssofzadeh et al., 2018), possibly because of bilateral sound and speech processing becoming more lateralized with age (Enge et al., 2020).

### Speech network brain-behaviour links

Reduced connectivity in the LiD group related most closely to the childrens’ listening skills, as reported by their caregivers, but also, more weakly, to objectively measured listening skills and cognition. Notably, speech-in-noise ability was not, in general, related to connectivity.

Structural and functional differences at the cortical level have not previously been directly compared with behavioral ability for children with LiD (e.g., Alvand et al., 2022, 2023; Farah et al., 2014; Pluta et al., 2014). This study’s brain-behavior relationships were restricted to the LiD and TD groups, and confirmed the specificity of the speech-listening deficit in children with LiD. The proportion of ROI-to-ROI connections having significant correlations with behavioral outcomes (across both the TD and LiD groups) increased from thalamic to primary auditory cortex, from primary to secondary auditory cortex and, finally, from anterior temporal to frontal cortex. This sequential progression of correlated neural activity mirrored the known route of speech synthesis and listening behavior. The ascending auditory system to primary auditory cortex is thought to process mainly foundational sound properties (e.g. pitch, Tramo et al., 2002). Secondary auditory cortex and posterior temporal areas process rhythm (Penhune et al., 1999), integrate across frequency, and perform initial phoneme encoding (DeWitt & Rauschecker, 2012). Higher-level, anterior temporal and frontal areas are involved in interpreting and comprehending speech (DeWitt & Rauschecker, 2012, 2013). More dynamic interplays at each of these levels has recently been illustrated with mice (e.g. Krall et al., 2024; Williamson & Polley, 2019), pointing a way to further, focused studies in humans.

No significant correlations were found across the Speech cortical network with spatial release from masking (SRM), a fundamental mechanism of auditory perception that aids listening in noisy environments (Litovsky, 2012). The behavioral measure indexing SRM was the Spatial Advantage score of the LiSN-S. This index has been reported to detect children having a specific, clinical Spatial Processing Disorder (SPD; Cameron et al., 2011; Cameron & Dillon, 2008). We chose the Spatial Advantage in part because it segregates sensory from cognitive aspects of listening (Moore & Dillon, 2018; Petley et al., 2021). In the baseline behavioral analysis in our longitudinal study, we did not find the Spatial Advantage to be a significant predictor of everyday listening skills, measured by the ECLiPS (Petley et al., 2021). However, it proved to be a significant predictor when using the greater power of the full longitudinal assessment, as discussed by Kojima and colleagues (2024). Cortical imaging results at the baseline time point, as presented here, were consistent with the behavioural findings of Petley et al. (2021); cortical connectivity showed low, mostly non-significant, correlations with the Spatial Advantage score. This supports the hypothesis that SPD may be a subcortical, predominantly sensory disorder that does not intersect, neurally or behaviorally, with caregiver reports of LiD. Such an interpretation is consistent with other behavioral data suggesting that SPD may be a distinct, easily treatable disorder of childhood (Cameron et al., 2012).

### Maturation of Speech and Sound functional connectivity

All groups showed considerable changes across the age range studied in both Speech and Sound networks, but not in the Visual network. We found age-related changes in connectivity in both the Speech and Sound networks in each of the participant groups between 6-14 years of age. Overall, these results fit with a long maturation of the auditory cortex into the teenage years (Calcus, 2024; Leibold, 2011; Litovsky, 2015) and beyond (Eggermont & Moore, 2011). This demonstrates that auditory experience, including language, contribute to the further maturation of the human auditory system, and associated cortical areas, despite the auditory system being functional around 25 weeks gestation (Graven & Browne, 2008) and showing primarily temporal activation to auditory stimuli during the third trimester (Loehn et al., 2026).

Functional connectivity changes in the Speech network of the TD children support the developmental framework of the central auditory nervous system set out by Eggermont and Moore (2011). Thalamocortical connections increase across the age groups (6/7 to 12/13/14 y.o.) and intracortical connections (between primary and secondary auditory cortices) strengthen until age 10/11 y.o. Similar increases in thalamocortical connectivity were observed in the TD children’s Sound network through to ages 12/13/14 y.o., reinforcing the generality of this developmental pattern across overlapping ROIs of the Speech and Sound network. This timeline aligns with the maturation of auditory spectral and temporal processing thresholds (7-9 years; (Moore et al., 2010), and the prolonged development of auditory stream segregation into adolescence (see Calcus, 2024 and Leibold, 2011 for reviews of non-linguistic behavioural and neurophysiological evidence).

Connectivity in later cortical stages of the auditory system (i.e., frontal areas) differed sharply between the clinical groups and TD children, especially in the youngest age group, suggesting the likelihood of more major differences at ages younger to those studied here. In contrast, the visual network as defined here was fully developed by age 6-7 across all groups and showed no age-related changes. These findings align with reports that auditory cortical maturation extends to age 20 (Eggermont & Moore, 2011), while visual development typically completes by age 7 (Mohammed & Khalil, 2020).

An unexpected drop in connectivity between auditory and fronto-temporal areas of the Speech network at age 12-13 in children with LiD may reflect a pre-existing weakness exacerbated by puberty, which has been proposed as an important auditory critical period (Pinheiro et al., 2024; Werker & Hensch, 2015). This decline may also be influenced by social shifts during the transition to secondary school (Norton, 1978), leading to the observed higher individual variability in auditory system development and differential impact across clinical groups (Kojima et al., 2024), with TD children relatively unaffected. This drop in connectivity may also reflect a bidirectional interaction between brain and behaviour during development whereby children with LiD have learnt to rely on non-speech strategies for processing the world as a consequence of both prior and ongoing difficulties with everyday listening (Jones & Rauwolf, 2025).

### Limitations

The activation likelihood estimation meta-analysis (Laird et al., 2011; Yarkoni et al., 2011) used to create our Speech, Sound and Visual networks (in neurosynth and GingerALE; Eickhoff et al., 2009; Turkeltaub et al., 2002; Yarkoni et al., 2011) did not distinguish between children and adults in the source studies, so we must assume they were a combination of both. This factor may account for some unexpected aspects of the ROIs used here. For example, in the Speech network, the right thalamus was not included and, while other speech ROIs were bilateral, they were not completely mirrored between hemispheres. Whatever the cause, this limitation is important to note, especially when interpreting lateralization and the group differences in connections between temporal and frontal areas in the Speech network.

Unfortunately, we do not have the same behavioural data with the ADHD group as we do for the LiD and TD groups. This would have allowed a more thorough examination of the brain-behaviour relationships between the two clinical groups. However, with the data we have available, clear conclusions can still be drawn with many new avenues for future research.

### Clinical implications

Here we provide neurobiological evidence for LiD as a distinct entity that is less predicted by objective measures than by holistic report. Simply by using a caregiver questionnaire regarding everyday listening difficulties, the data show us that cortical connectivity of a Sound network by children with LiD is similar to that of their TD peers. However, connectivity of a Speech specific network is clearly different. Thus we add to previous calls (e.g., Ertmer, 2011; Sanchez et al., 2022) for both speech intelligibility testing under challenging listening conditions (e.g., noise, reverberation) and caregiver report as standard parts of clinical pediatric auditory assessments, regardless of the referral route.

As evidenced by cross-sectional analysis across age groups, the differences in speech cortical connectivity continue into the teenage years for children with LiD, with little change. This adds to previous longitudinal behavioural analysis (Kojima et al., 2024) showing both cognitive and everyday listening difficulties persist in children with LiD as we followed them for up to six years. Together, these results suggest that childhood LiD is not a developmental delay.

Children with ADHD had a distinct auditory profile positioned between the TD children and children with LiD, with altered cortical connectivity for both speech and non-speech sounds. Both of the auditory networks continued to develop across age groups with different patterns of development for each group of children, thus adding to the growing transdiagnostic literature.

## Conclusions

We did not find a common etiology for LiD across diagnostic categories. Instead, cortical functional connectivity of children with LiD was characterised by a diminished Speech network relative to both TD children and those diagnosed with ADHD. Children with ADHD had weaker speech and sound auditory connectivity networks compared to TD children. Caregiver-reported everyday listening difficulties were more predictive of temporal neural connectivity across all children than were objective measures of hearing. Enhanced connectivity for the Speech and Sound networks was found across age in the TD group. However, children with either LiD or ADHD maintained relatively reduced neural connectivity across age groups. Current results, together with prior behavioral evidence, thus suggest that LiD is a higher level functional language disorder that is an enduring problem of neurodevelopment.

## Materials and Methods

Both studies were approved by the Institutional Review Board of Cincinnati Children’s Hospital (CCH) Research Foundation.

### Participants

Eighty-one children aged 6-14 took part in a larger listening study and were found to have normal hearing thresholds (< 25 dB HL at standard frequencies, Suppl. Figure 1). Of these children 39 were typically developing (TD) with no history or diagnosis of developmental disorders or delays and 42 were classified as having LiD from caregiver reports of everyday listening abilities (Figure 2; ECLiPS, Barry & Moore, 2021; Denys et al., 2024). Of these LiD children, 21 had a caregiver reported diagnosis of ADHD and 21 did not.

A further 23 children took part in a separate ADHD study (Figure 2). Fifteen of these children were diagnosed with ADHD after caregiver-administered semi-structured diagnostic interviews using the K-SADS (Kaufman et al., 1997). The remaining eight were classified as TD and were used to compare between study protocols (Suppl. Figure 7A). While it was not possible to conduct audiograms with the children from the ADHD study, each had a caregiver report of normal hearing and their medical histories were checked for audiology assessments, of which there were none.

### Resting state MRI acquisition

3 T Philips Ingenia scanners with a 65-channel head coil and Avotec audiovisual system were used for both the listening and ADHD studies. Both studies collected high-resolution T1-weighted anatomical scans with TR/TE=8.1/3.7 ms, FOV 25.6 x 25.6 x 16.0 cm, matrix 256 x 256 and slice thickness = 1mm. The listening study used a 5 minute long rs-fMRI acquisition with TR/TE = 2000/30 ms, voxel size = 2.5 × 2.5 × 3.5 mm, 39 axial slices in ascending slice order and 150 volumes. The ADHD study had a longer rs-fMRI protocol (10 minutes) with TR/TE = 2000/30 ms, voxel size = 2.8 × 2.8 × 3.0 mm, 39 axial slices in ascending slice order and 300 volumes. All participants were awake and non-sedated throughout their scanning protocols and were asked to look at a white fixation point in the centre of a black screen. In the ADHD study the children were asked to not take any ADHD relevant medication for 24 hours prior to the scan (Westbrook et al., 2020).

All the children were scanned at Cincinnati Children’s Hospital by the same MRI techs. However, different 3 T scanners were used for the two studies (TD/LiD and ADHD). See supplementary text for details comparing TD children from both studies to assess the influence of the different scanners on data collection (Suppl. Figure 7A).

### Resting state pre-processing

Pre-processing and analysis was performed in the CONN toolbox using standard spatial and temporal pipelines (Whitfield-Gabrieli & Nieto-Castanon, 2012). The ADHD scans were cut so that the first 150 volumes, matching the length of the listening study scans, were analyzed instead of the full 300. The Artifact Detection Tool (ART) within CONN was used to regress out framewise motion. A one-way ANOVA showed no group effect on the the number of frames regressed out (Table 1), *F*(3, 106) = 2.22, *p* = .09, η^2^_p_ = .059.

### Regions of interest selection and parcellation

Three networks were synthesized through large-scale activation likelihood estimation meta-analysis of published fMRI cortical activations by Neurosynth (Yarkoni et al., 2011) and GingerAle (Eickhoff et al., 2009; Turkeltaub et al., 2002). After applying an intensity threshold of 8 to the resulting activations from the meta-analyses the areas of activation were still too large for ROI-to-ROI rs-fMRI connectivity analysis. To make smaller more appropriate ROIs data-driven spatially constrained parcellation (Craddock et al., 2012) was applied to each network using the pediatric ADHD-200 sample (Bellec et al., 2017).

Neurosynth used 528 papers with the keyword ‘speech’ to create the Speech network which was parcellated into 49 ROIs (Figure 2C; Suppl. Figure 2). Similarly 603 papers with the keywords ‘vision’ and ‘visual system’ were parcellated into 22 ROIs for the Visual network (Figure 2C; Suppl. Figure 2). To ensure that the Sound network did not include fMRI activations from speech paradigms we used Neurosynth search terms such as ‘auditory’ and ‘listening’ (see Supp. Table 2 for full list of keywords) producing a list of 1,337 unique papers. We removed studies that used speech (syllables, words, multi-talker babble etc.) in their paradigms, leaving non-verbal stimuli (e.g., music, pure/complex tones, environmental sounds) paradigms. This left 195 papers whose cortical activations were entered into GingerAle and produced 69 parcellated ROIs (Figure 2B; Suppl. Figure 2). See Supp. Table 3 for ROI details (Brodman’s area, center of parcel coordinates and brain regions).

### Listening study questionnaires and behavioral tasks

*Listening skills* (ECLiPS; Barry & Moore, 2021; Denys et al., 2024) The children’s everyday listening and communication abilities were collected via the ECLiPS questionnaire . LiD group inclusion in the listening study was defined as an ECLiPS total scaled score < 7 or a previous diagnosis of APD (n = 11).

The ECLiPS is composed of 38 items describing commonly observed behaviors (e.g., the ability to follow conversations) which the caregivers rate how much they agreed to each statement on a five-point Likert scale from strongly disagree to strongly agree. Responses are averaged into five subscales (speech & auditory processing; environmental & auditory sensitivity; language/ literacy/ laterality; memory & attention; and pragmatic & social skills), three composite scores (Language, Listening and Social) or a total composite score. All of the ECLiPS scores are scaled for age with the composite and total scores standardized for a population mean of 10 (SD = 3), based on British data (Barry et al., 2015).

*Communication skills* (CCC-2; Bishop, 1998) Caregivers also responded to 70 items in the CCC-2 questionnaire asking about their child’s communication skills. Responses make up 10 subscales (speech, syntax, semantics, coherence, initiation, scripted language, context, nonverbal communication, social relations and interests). These subscales then combine to create two composite scores, a General Communication Composite (GCC, all subscales apart from the social relations and interests) and Social Interaction Differences Index. We used the GCC in our analysis.

*Listening skills* (SCAN-3:C; Keith, 2000) This is a standardized test for APD for children aged 5 to 12 years. We administered the SCAN-3:C either in a sound-attenuated booth or a quiet room via a laptop with MAYA USB soundcard and Sennheiser HD 215 headphones. The standardized composite score was used in analysis.

The SCAN-3:C is made up of four subtests where participants repeat back: low-pass filtered, monosyllabic words in quiet (low-pass Filtered Words); unfiltered monosyllabic words against multi-talker speech at +8dB SNR (Auditory Figure Ground); monosyllabic words (Competing Words-directed ear) or sentences (Competing Sentences) presented in either the left or right ear while different monosyllabic words or sentences (respectively) are presented simultaneously in the other ear. Subtest scores are age-scaled and are combined to make a standardized composite score.

*Speech Hearing in Noise* (LiSN-S; Cameron & Dillon, 2007) The Listening in Spatialized Noise-Sentences (LiSN-S) test measures speech listening in informational masking. We administered the US version of the LISN-S (Cameron et al., 2009) either in a sound-attenuated booth or a quiet room via a laptop with task-specific soundcard and Sennheiser HD 215 headphones. We used the standardized Spatial advantage score in our analysis.

In the LISN-S participants are asked to repeat back target sentences (T) while ignoring two sets of distractor sentences (D1, D2) in four different listening conditions. The talkers may have the same or different voices, or come from the same or different directions (0°, ± 90° azimuth). From the four listening conditions, the LISN-S software calculates three difference scores to give isolation between the auditory and cognitive contributions of the task (Moore & Dillon, 2018): Talker advantage (different voices - same voice); Spatial advantage (different directions - same direction); and Total advantage (different voices and directions - same voices and directions).

*Cognition* (National Institutes of Health (NIH) Toolbox Cognition Battery; Weintraub et al., 2013) We administered the NIH Toolbox either in a sound-attenuated booth or a quiet room via online or an iPad app following the NIH Toolbox procedures. We used the Early Childhood Composite score in our analysis. This composite is an average of four standardized cognitive task scores: Picture Vocabulary test (PVT); Flanker test (Flanker); Dimensional Change Card Sort test (DCCS); and Picture Sequence Memory test (PSMT). The PVT tests vocabulary knowledge through an adaptive assessment where the participant matches the audio recording of a word with one of four pictures that most closely matches the meaning of the word. The Flanker, tests selective attention and the executive control ability of inhibition. The task asks participants to report the direction of a central target (an arrow or fish dependent on the participant’s age) in a horizontal string of four other distractors (also arrow or fish) that may be congruent (same direction as the target) or incongruent (opposite direction). The DCCS assesses attention switching by asking participants to switch between two rules for sorting cards trial by trial (e.g., color and then by shape). Finally, the PSMT assesses episodic memory by first telling the participant audio-recorded stories with an increasing sequence of cards (6 to 18, depending on age) to illustrate each story, and then asking the participant to recall the story by sorting the now jumbled illustrative cards into the correct order.

### Analysis

The mean time course of all voxels within each ROI was used to calculate individual pairwise Pearson correlations for ROI-to-ROI analysis within each network (Speech, Sound and Visual). The *r* values were normalized to *z* values via Fisher’s z-transformation. Statistical thresholds were set to *p* < .05 (FDR-corrected) at the single voxel level, and the resulting connections were thresholded at seed-level by intensity with FDR correction (*p* < .05).

#### Within network connectivity: Between groups

Connectivity between ROIs for each network (Speech, Sound and Visual) were first compared between groups using a general linear model (GLM) while controlling for age. Post hoc GLMs were run between groups for networks with significant group level differences. Second, the averaged within-network functional connectivity values for each network were extracted from CONN and compared between groups using ANOVA and post-hoc t-tests with Holm-correction in JASP.

GLMs also compared TD children from the listening and ADHD studies as a data quality check; children with LiD with and without an APD diagnosis; and children with LiD with and without a speech, language pathology referral.

#### Relating Speech network connectivity to behavior

Corrected t-tests compared the strength of connectivity (z-score) of individual Speech network ROI-to-ROI pairs connectivity (z-score) between groups (TD and LiD) (Figure 4). Only these two groups were used in this analysis due to the depth of behavioural measures and audiological assessments used in the listening study. ROI pairs were grouped into basic auditory processing to primary auditory cortex; primary to secondary auditory cortex; and temporal-to-frontal.

Connections with significant group differences were further explored using planned Pearson correlations with Bonferroni correction to assess brain-behavior relationships (Figure 5; Suppl. Figure 8). Behavioral outcomes were listening skills (subjective: ECLiPS total score; and objective: SCAN composite) communication (GCC from the CCC-2), speech in noise skills (LiSN-S spatial advantage score) and cognition (NIH ECC).

#### Within network connectivity: Across development

GLMs were used to compare age groups within each diagnostic group (TD, LiD and ADHD). Post hoc GLMs were run between age groups for networks with significant overall age group differences.

## Data Availability

ROI files can be found at osf.io/qzkh4

## Acknowledgements

Thanks to Audrey Perdew and Nicholette Sloat for recruitment and testing in the listening study; to Dr Kimberly Leikin and Dr Ronan McGarrigle for assisting in the listening study MRI acquisition; to Amanda Withrow, Dana Schindler, and Josalyn Foster for recruitment and testing in the ADHD study; to the CCH MRI techs Matt Lanier, Lacey Haas, Kaley Bridgewater, Kelsey Murphy, Brynne Williams and Marty Jones for MRI acquisitions in both studies; Megan Griffiths for Figure 5 and Supp. Figure 8; and Dr Chelsea Blankenship and Jody Caldwell-Kurtzman for their continued support. Many thanks to Dr Jon Dudley for analysis discussions.

## Funding

This research was supported by NIH R01DC014078, NIH K23MH108603, and a Cincinnati Children’s Research Foundation Trustee Award. David Moore was part supported by the UK National Institute for Health and Care Research (NIHR) Manchester Biomedical Research Centre (BRC) (NIHR203308). Hannah Stewart was part supported by UKRI Future Leaders Fellowship MR/X035999/1.

## Competing interests

All authors report no conflicts of interest.

## Data and materials availability

ROI files can be found at osf.io/qzkh4.

## Author contributions

HJS - meta-analysis design and analysis, MRI acquisition and analysis, figures and tables, manuscript preparation and revisions

EC - meta-analysis analysis, figures, assisted with manuscript preparation and revisions LLH - study design, manuscript preparation and revisions

JEP - analysis and presentation, manuscript preparation and revisions LT - ADHD study, manuscript revisions

SPB - ADHD study, manuscript revisions

JV - study design, analysis and presentation, manuscript preparation

DRM - meta-analysis design, study design and presentation, analysis and presentation, manuscript preparation and revisions

## Abbreviations

ADHD: attention-deficit/hyperactivity disorder
APD: auditory processing disorder
BOLD: blood oxygenation level dependent
DMN: default mode network
FDR: false discovery rate
FOV: field-of-view
IC: insular cortex
IPS: intraparietal sulcus
LiD: listening difficulties
MRI: magnetic resonance imaging
MTG: middle temporal gyrus
ROI: region of interest
rsMRI: resting state MRI
STG: superior temporal gyrus
TD: typically developing
TE: time to echo
TFC: temporal fusiform cortex
TL: temporal lobe
TR: repetition time

## Notes

### Competing Interest Statement

The authors have declared no competing interest.

### Author Declarations

IRB of Cincinnati Children's Hospital Medical Centre gave ethical approval for this work.

### Summary of Updates

Rewrite throughout and new significance statement.

