## Supplementary material for "Forebrain speech networks specifically impaired in children with listening difficulties": Supplmental material

**Supplementary Figure 1: All the children from the Listening study had ”normal hearing” as defined by pure tone audiometry (≤ 20 dB PTA for frequencies 0.25 - 8 kHz bilaterally).** The vertical dashed line shows the start of extended high frequencies (10, 12.5, 14 and 16 kHz).

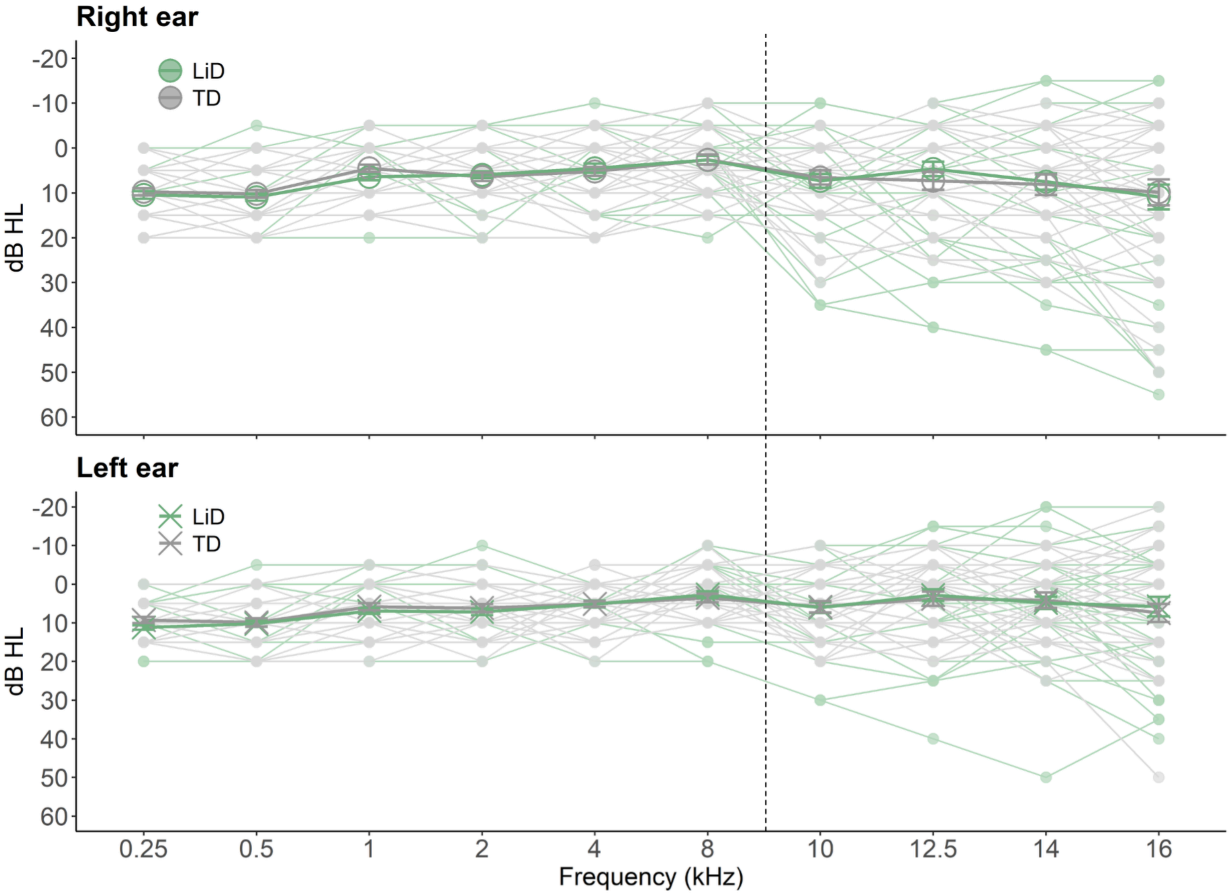

**Supplementary Table 1**: **Group differences in averaged connectivity from across the Speech and Sound networks.** Significant group differences in averaged connectivity from across the Speech and Sound networks. Post-hoc results from the ANOVA in Table 1, see also Fig. 3D.

| Network | Group | | df | t | p | Cohen's d |
|---|---|---|---|---|---|---|
| Speech | LiD | TD | 79 | -11.94 | < .001 | .35 |
| | ADHD | TD | 60 | -6.79 | < .001 | .29 |
| | | LiD | 63 | 3.40 | < .001 | .46 |
| Sound | LiD | TD | 79 | -0.24 | .81 | .22 |
| | ADHD | TD | 60 | -2.53 | .039 | .28 |
| | | LiD | 63 | 2.36 | .040 | .28 |

**Supplementary Table 2**: **The search terms used in Neurosynth to create the Sound network.** We removed studies that reported activation to speech stimuli, leaving 195 unique papers ready for activation likelihood estimation meta-analysis using GingerALE.

| Neuorysnth search term | # studies (some overlap between search terms) |
|---|---|
| Auditory | 1055 |
| Audio | 60 |
| Listening | 204 |
| Listeners | 60 |
| Listened | 109 |
| Hearing | 104 |
| Tone | 97 |
| Complex tone | 2 |
| Hear | 58 |
| Listen | 46 |
| Listening | 43 |

**Supplementary Figure 2: Networks created by regions of interest (ROI) as defined by Neurosynth with search terms of "Speech", non-speech "Sound" (see Supp. Table 1 for more details), and a "Visual" control.** Parcellations of these ROIs by the Craddock ADHD 200 atlas can be seen in Figure 1 C, D and E. Full ROI details are in Supplementary table 2.

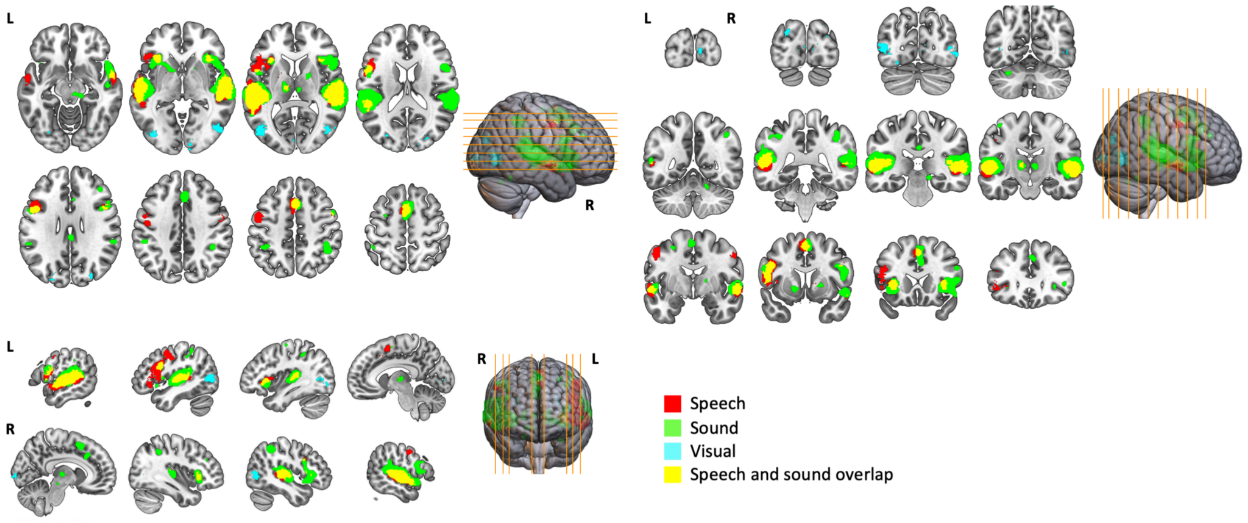

**Supplementary Table 3. MNI coordinates and Brodmann area (BA) for the ROIs as defined by parcellating the "Speech", "Sound" and "Visual" networks with the Craddock ADHD 200 atlas.** See Figure 1C for a visualization of these ROIs, and Supplementary Figure 2 for each network before parcellation. Cluster threshold = 4.0 voxels, p-FDR = .95. BA and brain region area have been left blank when no labelling was available for the ROI coordinates using the Harvard-Oxford cortical and subcortical structural atlases via FSLeyes.

| Network | ROI | BA | MNI ROI coords Center of parcel | | | Brain Regions (Harvard-Oxford atlases) |
|---|---|---|---|---|---|---|
| | | | x | y | z | |
| **Speech** | 7 | 48 | 48 | -14 | 6 | R Heschl's gyrus |
| | 8 | | -22 | 2 | 2 | L Putamen |
| | 10 | 21 | 64 | -24 | 2 | R superior temporal gyrus |
| | 13 | 32 | 0 | 18 | 48 | L paracingulate gyrus |
| | 22 | 22 | 60 | -6 | -8 | R superior temporal gyrus |
| | 27 | 45 | -46 | 30 | 14 | L inferior frontal gyrus |
| | 31 | | 12 | 8 | 4 | R cerebral white matter |
| | 32 | 22 | -60 | -24 | 8 | L planum temporale |
| | 35 | 6 | 2 | 2 | 64 | R supplementary motor cortex |
| | 36 | 22 | -58 | -6 | -8 | L superior temporal gyrus |
| | 48 | 47 | -48 | 26 | -6 | L frontal orbital cortex |
| | 54 | | -20 | 4 | 4 | L putamen |
| | 59 | 47 | 36 | 22 | 2 | R insular cortex |
| | 60 | 4 | 50 | -10 | 40 | R precentral gyrus |
| | 66 | 48 | -48 | 0 | 10 | L pars opercularis |
| | 67 | | -10 | -16 | 6 | L thalamus |
| | 69 | 19 | 24 | -60 | -22 | |
| | 70 | 22 | 52 | -28 | 2 | R superior temporal gyrus |
| | 73 | 48 | -46 | 20 | 28 | L middle frontal gyrus |

| | | | | | |
|---|---|---|---|---|---|
| 83 | 4 | -48 | -12 | 40 | L precentral gyrus |
| 87 | 6 | -50 | -4 | 46 | L precentral gyrus |
| 89 | 32 | 0 | 10 | 48 | L paracingulate gyrus |
| 94 | 6 | 52 | 0 | 44 | R precentral gyrus |
| 106 | 22 | 60 | -38 | 8 | R supramarginal gyrus |
| 109 | 22 | -58 | -44 | 10 | L supramarginal gyrus |
| 110 | 38 | 54 | 12 | -20 | R temporal pole |
| 113 | | -12 | -18 | 6 | L thalamus |
| 114 | 4 | -54 | -2 | 32 | L precentral gyrus |
| 117 | 22 | 58 | -12 | 2 | R planum temporale |
| 122 | 22 | -62 | -18 | -4 | L superior temporal gyrus |
| 124 | 47 | -36 | 22 | 2 | L insular cortex |
| 129 | 48 | 54 | -2 | 10 | R central opercular cortex |
| 136 | 22 | 58 | -24 | 10 | R planum temporale |
| 138 | 48 | 50 | -10 | -2 | R Heschl's gyrus |
| 144 | 48 | -50 | -18 | 2 | L Heschl's gyrus |
| 148 | 6 | -4 | 8 | 60 | L supplementary motor cortex |
| 155 | 38 | 56 | 6 | -6 | R temporal pole |
| 157 | 48 | -56 | 2 | 4 | L pars opercularis |
| 160 | 6 | -2 | 4 | 58 | L supplementary motor cortex |
| 168 | 38 | -48 | 22 | -6 | L pars triangularis |
| 173 | 48 | 46 | 12 | 28 | R inferior frontal gyrus |
| 175 | 44 | -46 | 10 | 28 | L inferior frontal gyrus |
| 176 | 41 | -46 | -32 | 12 | L planum temporale |
| 178 | 7 | -28 | -60 | 44 | L lateral occipital cortex |
| 179 | 48 | -50 | 16 | 16 | L pars opercularis |
| 185 | 48 | 44 | -24 | 10 | R Heschl's gyrus |

| | | | | | | |
|---|---|---|---|---|---|---|
| | 188 | 22 | -52 | -46 | 12 | L supramarginal gyrus |
| | 197 | 22 | -56 | -36 | 2 | L superior temporal gyrus |
| | 200 | 48 | 50 | 14 | 26 | R inferior frontal gyrus |
| **Sound** | 4 | 45 | 44 | 34 | 18 | R middle frontal gyrus |
| | 7 | 48 | 44 | -14 | 4 | R Heschl's gyrus |
| | 8 | | -16 | -4 | 0 | L pallidum |
| | 10 | 21 | 66 | -26 | 4 | R superior temporal gyrus |
| | 13 | | 2 | 20 | 46 | R paracingulate gyrus |
| | 22 | 22 | 58 | -6 | -8 | R superior temporal gyrus |
| | 31 | | 18 | 8 | 22 | R caudate |
| | 32 | 22 | -62 | -26 | 12 | L planum temporale |
| | 33 | | -14 | -10 | 8 | L thalamus |
| | 35 | 6 | 4 | 10 | 60 | R superior frontal gyrus |
| | 36 | 22 | -56 | -6 | -8 | L superior temporal gyrus |
| | 39 | 47 | 46 | 28 | 0 | R inferior frontal gyrus |
| | 42 | 48 | 60 | -38 | 26 | R supramarginal gyrus |
| | 45 | | 20 | 2 | 4 | R palidum |
| | 51 | 40 | -54 | -36 | 40 | L supramarginal gyrus |
| | 53 | 40 | 42 | -44 | 44 | R supramarginal gyrus |
| | 54 | 11 | -20 | 12 | -2 | L putamen |
| | 56 | 6 | -4 | -2 | 66 | L supplementary motor cortex |
| | 58 | 6 | 10 | 10 | 54 | R paracingulate gyrus |
| | 59 | 47 | 36 | 22 | 0 | R insular cortex |

| | | | | | |
|---|---|---|---|---|---|
| 66 | 48 | -46 | -10 | 10 | L central opercular cortex |
| 67 | | -4 | -16 | 2 | L thalamus |
| 69 | 37 | 18 | -54 | -22 | |
| 70 | 48 | 50 | -26 | 2 | R superior temporal gyrus |
| 74 | 45 | 42 | 34 | 30 | R middle frontal gyrus |
| 80 | 23 | 2 | -32 | 28 | R cingulate gyrus |
| 83 | 40 | -48 | -34 | 42 | L supramarginal gyrus |
| 89 | 32 | 4 | 14 | 46 | R paracingulate gyrus |
| 94 | 6 | 52 | 4 | 46 | R precentral gyrus |
| 97 | 6 | -28 | -4 | 54 | L middle frontal gyrus |
| 106 | 42 | 62 | -42 | 14 | R supramarginal gyrus |
| 107 | 23 | 2 | -28 | 30 | R cingulate gyrus |
| 109 | 22 | -60 | -44 | 14 | L supramarginal gyrus |
| 112 | 6 | 8 | 18 | 54 | R superior frontal gyrus |
| 113 | | -14 | -18 | 8 | L thalamus |
| 114 | 6 | -54 | 2 | 24 | L precentral gyrus |
| 117 | 22 | 60 | -10 | 6 | R planum temporale |
| 120 | 24 | 4 | 24 | 32 | R cingulate gyrus |
| 122 | 22 | -62 | -18 | -2 | L superior temporal gyrus |
| 124 | 47 | -34 | 20 | 0 | L insular cortex |
| 125 | | 18 | 0 | 12 | R cerebral white matter |
| 129 | 6 | 56 | 8 | 24 | R precentral gyrus |
| 130 | 25 | -12 | 6 | -6 | L pallidum |
| 136 | 42 | 62 | -28 | 18 | R planum temporale |

| | | | | | |
|---|---|---|---|---|---|
| 137 | 6 | -38 | -14 | 62 | L precentral gyrus |
| 138 | 48 | 48 | -10 | -4 | R planum polare |
| 144 | 48 | -46 | -18 | 2 | L Heschl's gyrus |
| 148 | 6 | -2 | 12 | 60 | L superior frontal gyrus |
| 149 | 40 | 46 | -46 | 48 | R supramarginal gyrus |
| 155 | | 52 | 12 | -4 | R temporal pole |
| 156 | 48 | -58 | -40 | 26 | L parietal operculum cortex |
| 157 | 48 | -54 | -2 | 4 | L central opercular cortex |
| 160 | | 0 | 2 | 58 | L supplementary motor cortex |
| 162 | 42 | 52 | -40 | 16 | R supramarginal gyrus |
| 166 | 40 | -46 | -44 | 52 | L supramarginal gyrus |
| 168 | 38 | -44 | 16 | -6 | L insular cortex |
| 172 | 4 | -42 | -14 | 60 | L precentral gyrus |
| 173 | 44 | 46 | 10 | 26 | R precentral gyrus |
| 174 | | 2 | -22 | -8 | |
| 175 | 44 | -46 | 8 | 26 | L inferior frontal gyrus |
| 176 | 41 | -44 | -32 | 14 | L parietal operculum cortex |
| 178 | 40 | -38 | -42 | 46 | L superior parietal lobule |
| 179 | 44 | -52 | 10 | 20 | L inferior frontal gyrus |
| 183 | | -30 | -64 | -28 | |
| 184 | 30 | 16 | -26 | -10 | |
| 185 | 48 | 46 | -24 | 12 | R Heschl's gyrus |
| 188 | 21 | -50 | -46 | 16 | L supramarginal gyrus |

| | | | | | | |
|---|---|---|---|---|---|---|
| | 197 | 21 | -56 | -34 | 2 | L superior temporal gyrus |
| | 200 | 48 | 52 | 16 | 16 | R inferior frontal gyrus |
| **Visual** | 28 | 19 | -20 | -86 | 34 | L lateral occipital cortex |
| | 29 | 17 | 10 | -96 | -2 | R occipital pole |
| | 37 | | 2 | -84 | 4 | R supracalcarine cortex |
| | 55 | 19 | -28 | -76 | -14 | L occipital fusiform gyrus |
| | 63 | 19 | -22 | -78 | 26 | L lateral occipital cortex |
| | 64 | 19 | 28 | -82 | 26 | R lateral occipital cortex |
| | 76 | 17 | 12 | -94 | 8 | R occipital pole |
| | 84 | 19 | 28 | -82 | 16 | R lateral occipital cortex |
| | 93 | 19 | -36 | -82 | -8 | L lateral occipital cortex |
| | 96 | 17 | -6 | -94 | 2 | L occiptal pole |
| | 119 | 19 | 30 | -72 | -12 | R occipital fusiform gyrus |
| | 131 | 19 | -26 | -84 | 24 | L lateral occipital cortex |
| | 134 | 37 | 46 | -72 | 4 | R lateral occipital cortex |
| | 141 | 37 | -48 | -64 | -8 | L lateral occipital cortex |
| | 142 | 19 | 48 | -74 | -4 | R lateral occipital cortex |
| | 146 | 19 | 24 | -88 | 38 | R occipital pole |
| | 161 | | 30 | -64 | 62 | R lateral occipital cortex |
| | 171 | 19 | -46 | -76 | 6 | L lateral occipital cortex |
| | 180 | 17 | -10 | -98 | 12 | L occipital pole |
| | 187 | 37 | 50 | -70 | -4 | R lateral occipital cortex |

| | | | | | |
|---|---|---|---|---|---|
| 188 | 37 | -42 | -64 | 6 | L lateral occipital cortex |
| 190 | 19 | -36 | -80 | -10 | L occipital fusiform gyrus |

**Supplementary Figure 3: A single significant different ROI-ROI pair between the ADHD and LiD groups in the Visual network.** ROI-ROI resting state connectivity where thicker, more saturated color lines represent stronger connections between cortical areas. Anatomical representations of these ROI networks in full brain and brain slices are presented in Suppl. Figure 2. Heat maps further showing the quantitative strength of these connections can be found in Suppl. Figure 5. Glass brain images are in neurological convention and were created in CONN functional connectivity toolbox (https://web.conn-toolbox.org).

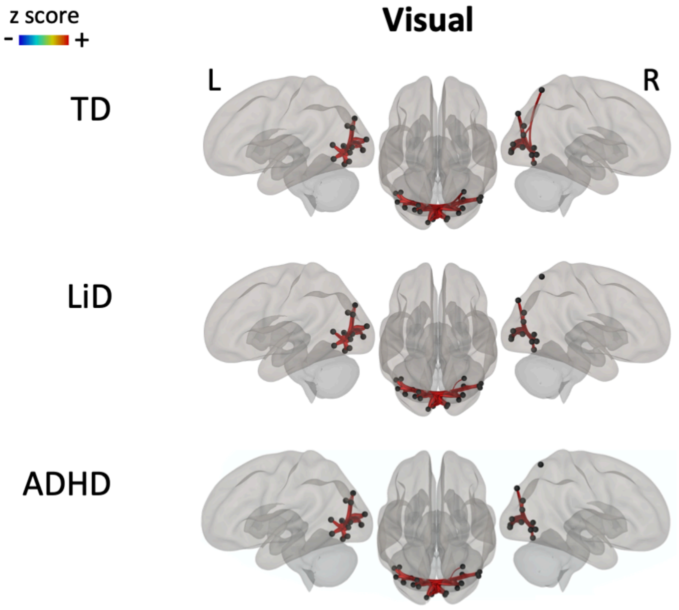

**Supplementary Figure 4**: **Highly specific differences in averaged connectivity across the A) Speech network between groups, but not in the B) Sound or C) Visual networks.** Averaged connectivity of each network across all of its ROI-to-ROI-pairs, separated by group (TD grey, LiD green, ADHD blue; see also Table 1). Boxplots show the groups' upper quartile, median and lower quartile.

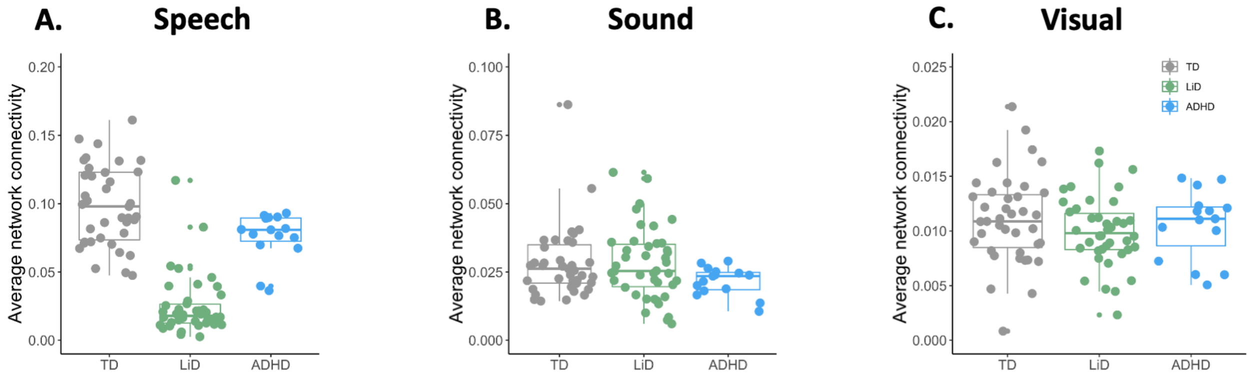

**Supplementary Figure 5: Heat maps showing the strength of the ROI-ROI resting state connectivity per group as shown in Figure 2 for (A) Speech, (B) Sound and (C) Visual.** The color scale for connectivity is consistent across all graphs.

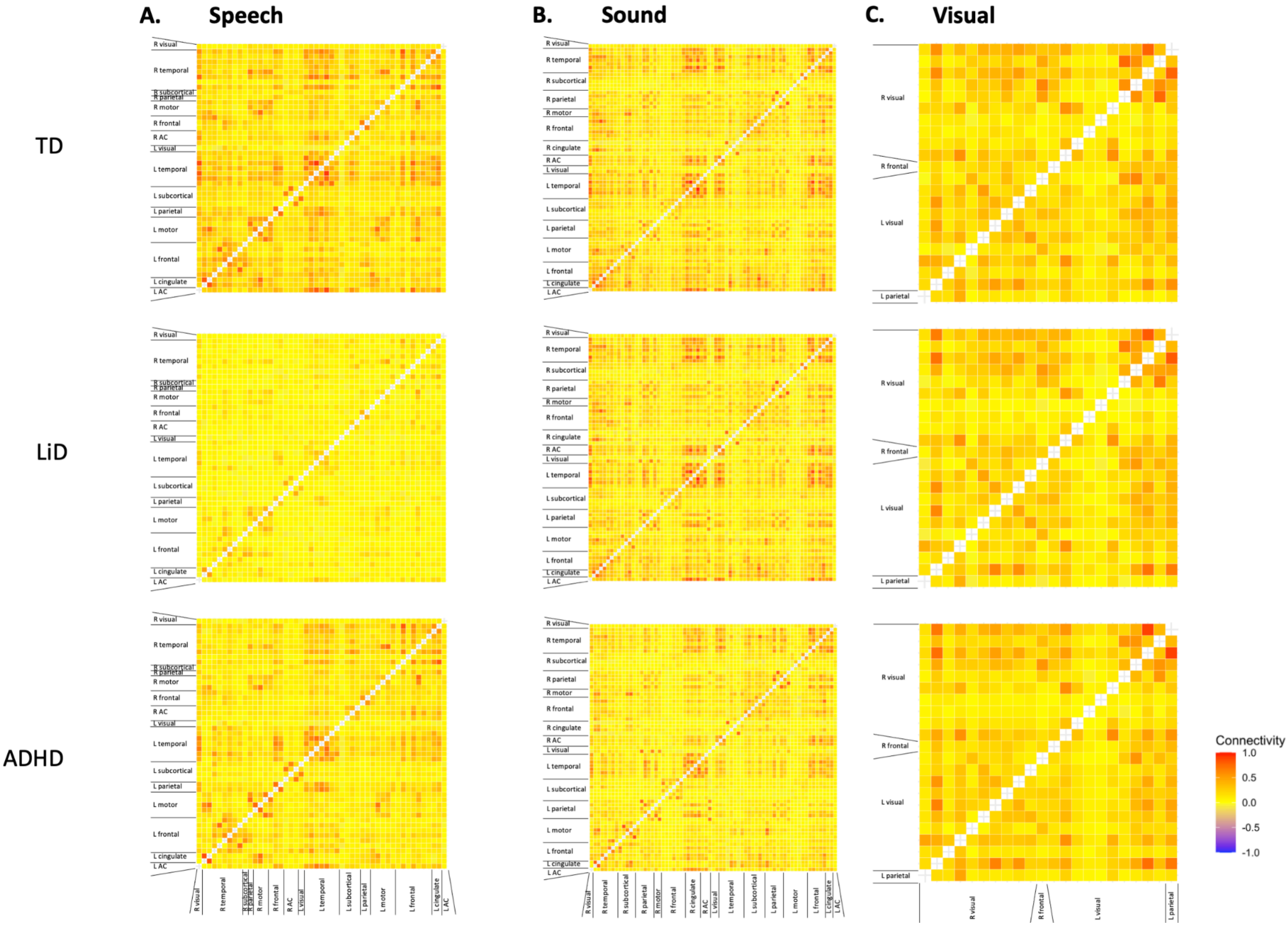

**Supplementary Figure 6: Large significant group differences (without the effect of age) were found in the (A) Speech network, especially between TD children and children with LiD.** Minor group differences were found in the (B) Sound and (C) Visual networks. Thicker, more saturated color lines represent stronger differences in connectivity between ROIs.

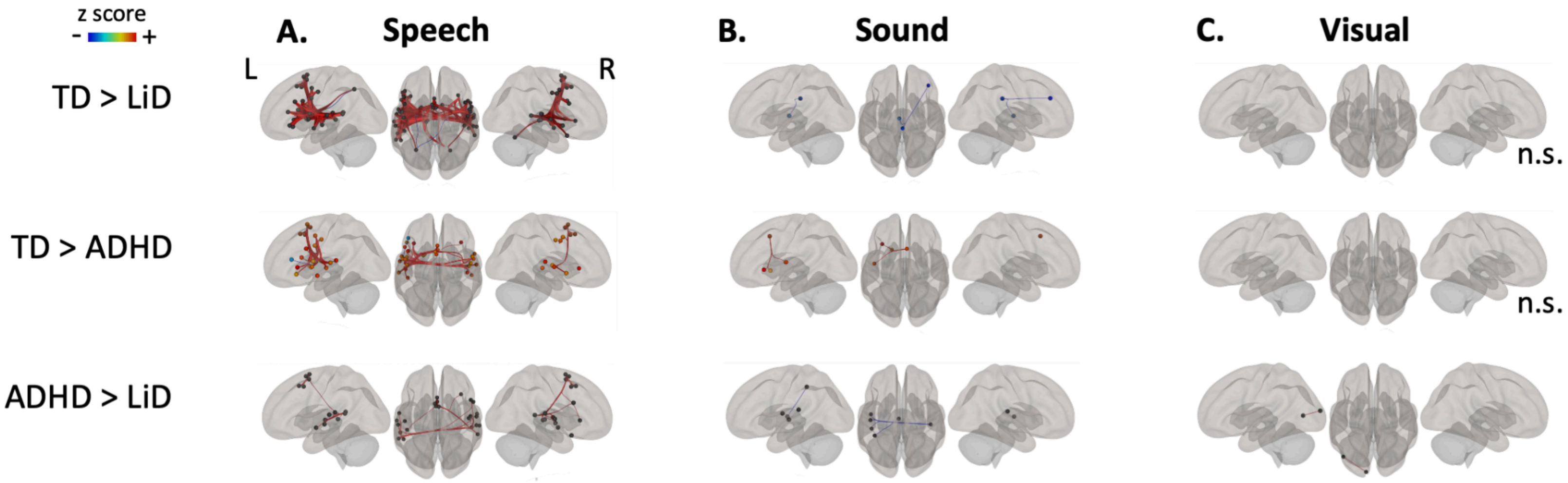

**Supplementary Figure 7: Minor differences between subsets of the children between and within the Listening and ADHD studies.** Subsets are A) TD children selected from the listening study to be age and gender matches to the TD children of the ADHD study; B) children with LiD with and without a formal APD diagnosis; and C) children with LiD who had and had not been referred to speech language pathology.

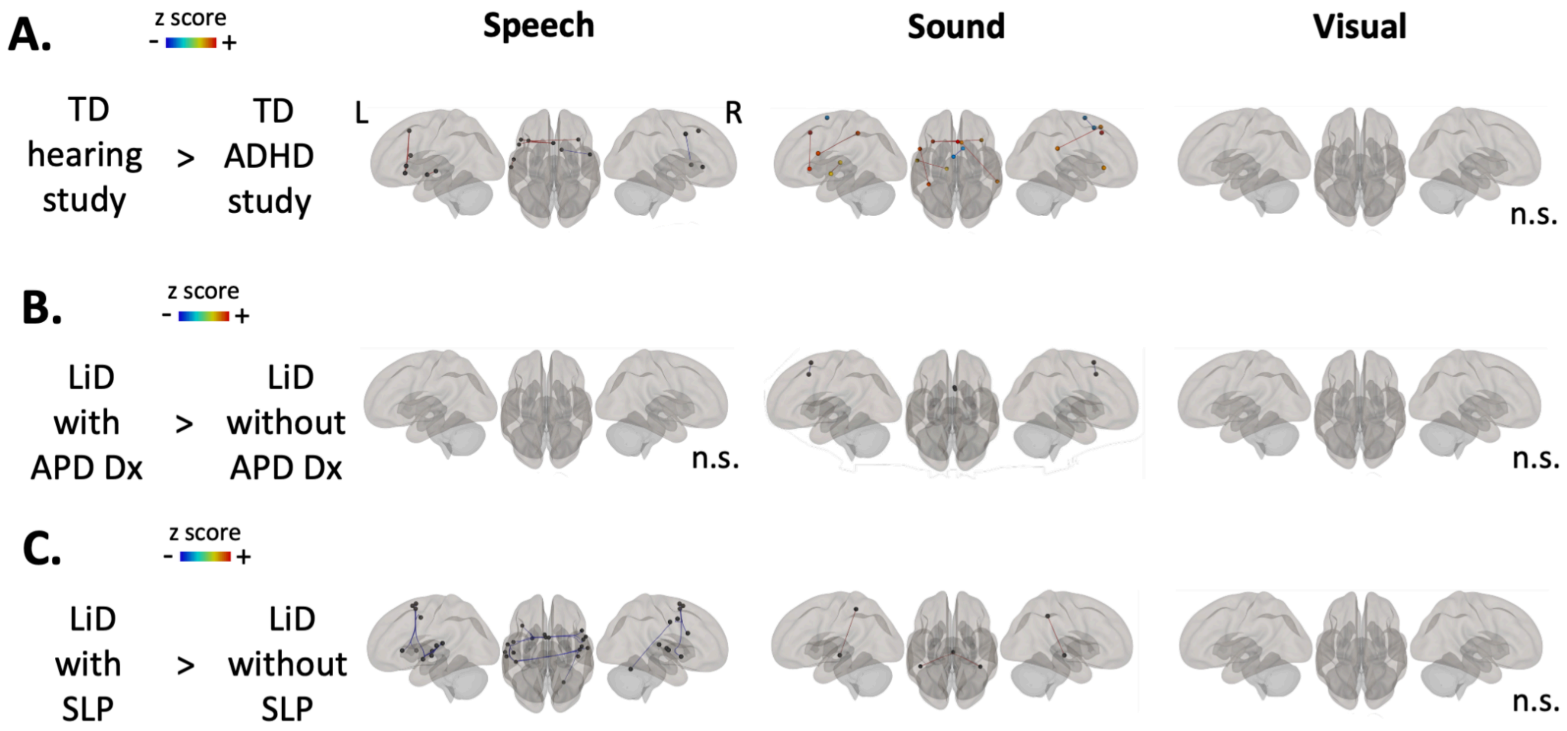

**Supplementary Table 4**: **ROI-to-ROI pairs used in the brain-behavior analysis.** Independent t-tests (df = 76) compared groups on connectivity strength between ROI pairs in: the thalamus and primary auditory cortex (left thalamus and bilateral Heschl's gyrus); primary and secondary auditory cortical areas (bilateral Heschl's gyrus and temporal gyrus); and secondary auditory and temporo-frontal cortical areas (bilateral temporal gyrus and left pars triangularis, frontal orbital cortex, IC and pars opercularis). This is illustrated in Figure 3. ROI pairs with a significant group difference that survived family correction (8, 16 and 30, respectively) are highlighted in blue and were used in the brain-behavior analysis in Figure 4. ROI details are in Supp. Table 2.

| Grouping | ROI - ROI | t | p | Cohen's d |
|---|---|---|---|---|
| Thalamus → Primary AC | 67-144 | 2.969 | 0.004 | 0.672 |
| | 67-176 | 3.202 | 0.002 | 0.725 |
| | 67-7 | 1.301 | 0.197 | 0.295 |
| | 67-185 | 1.911 | 0.06 | 0.433 |
| | 113-144 | 1.766 | 0.081 | 0.4 |
| | 113-176 | 2.774 | 0.007 | 0.628 |
| | 113-7 | 1.131 | 0.262 | 0.256 |
| | 113-185 | 1.983 | 0.051 | 0.449 |
| Primary AC → Secondary AC | 144-109 | 6.24 | < .001 | 1.413 |
| | 144-197 | 4.879 | < .001 | 1.105 |
| | 144-22 | 3.171 | 0.002 | 0.718 |
| | 144-70 | 5.784 | < .001 | 1.31 |
| | 176-109 | 6.311 | < .001 | 1.412 |
| | 176-197 | 2.677 | 0.009 | 0.606 |
| | 176-22 | 2.276 | 0.026 | 0.515 |
| | 176-70 | 4.637 | < .001 | 1.05 |
| | 7-109 | 3.036 | 0.003 | 0.688 |
| | 7-197 | 1.762 | 0.082 | 0.399 |
| | 7-22 | 1.018 | 0.312 | 0.231 |
| | 7-70 | 2.655 | 0.01 | 0.601 |
| | 185-109 | 3.078 | 0.003 | 0.697 |
| | 185-197 | 4.277 | < .001 | 0.969 |
| | 185-22 | 2.55 | 0.013 | 0.577 |
| | 185-70 | 2.195 | 0.031 | 0.497 |

| | | | | |
|---|---|---|---|---|
| Secondary AC → Left frontal | 168-144 | 2.961 | 0.004 | 0.671 |
| | 168-197 | 7.486 | < .001 | 1.695 |
| | 168-109 | 5.3 | < .001 | 1.2 |
| | 168-22 | 3.582 | < .001 | 0.811 |
| | 168-70 | 4.644 | < .001 | 1.052 |
| | 48-144 | 3.086 | 0.003 | 0.699 |
| | 48-197 | 7.296 | < .001 | 1.652 |
| | 48-109 | 6.05 | < .001 | 1.37 |
| | 48-22 | 4.506 | < .001 | 1.02 |
| | 48-70 | 4.343 | < .001 | 0.983 |
| | 124-144 | 3.823 | < .001 | 0.866 |
| | 124-197 | 4.628 | < .001 | 1.048 |
| | 124-109 | 6.912 | < .001 | 1.565 |
| | 124-22 | 1.061 | 0.292 | 0.24 |
| | 124-70 | 4.696 | < .001 | 1.063 |
| | 157-144 | 10.031 | < .001 | 2.272 |
| | 157-197 | 6.001 | < .001 | 1.359 |
| | 157-109 | 5.401 | < .001 | 1.215 |
| | 157-22 | 4.035 | < .001 | 0.914 |
| | 157-70 | 6.937 | < .001 | 1.571 |
| | 66-144 | 5.105 | < .001 | 1.156 |
| | 66-197 | 1.928 | 0.058 | 0.437 |
| | 66-109 | 4.521 | < .001 | 1.024 |
| | 66-22 | 1.346 | 0.182 | 0.305 |
| | 66-70 | 3.195 | 0.002 | 0.723 |
| | 179-144 | 4.295 | < .001 | 0.973 |
| | 179-197 | 6.878 | < .001 | 1.558 |
| | 179-109 | 5.315 | < .001 | 1.189 |
| | 179-22 | 3.667 | < .001 | 0.83 |
| | 179-70 | 5.686 | < .001 | 1.288 |
| Secondary AC → Right frontal | 144-60 | 4.504 | < .001 | 1.02 |
| | 197-60 | 1.867 | 0.066 | 0.423 |

| | | | |
|---|---|---|---|
| 109-60 | 2.023 | 0.046 | 0.453 |
| 22-60 | 0.98 | 0.33 | 0.222 |
| 70-60 | 3.033 | 0.003 | 0.687 |
| 144-94 | 5.261 | < .001 | 1.191 |
| 197-94 | 3.316 | 0.001 | 0.751 |
| 109-94 | 1.628 | 0.105 | 0.365 |
| 22-94 | 2.619 | 0.011 | 0.593 |
| 70-94 | 4.196 | < .001 | 0.95 |
| 144-59 | 3.624 | < .001 | 0.821 |
| 197-59 | 1.482 | 0.143 | 0.336 |
| 109-59 | 2.97 | 0.004 | 0.673 |
| 22-59 | 0.899 | 0.371 | 0.204 |
| 70-59 | 1.857 | 0.067 | 0.421 |
| 144-129 | 5.392 | < .001 | 1.221 |
| 197-129 | 3.861 | < .001 | 0.874 |
| 109-129 | 2.667 | 0.009 | 0.596 |
| 22-129 | 1.165 | 0.248 | 0.264 |
| 70-129 | 2.406 | 0.019 | 0.545 |
| 144-110 | 1.293 | 0.201 | 0.299 |
| 197-110 | 4.328 | < .001 | 0.996 |
| 109-110 | 3.112 | 0.003 | 0.715 |
| 22-110 | 2.622 | 0.011 | 0.603 |
| 70-110 | 3.943 | < .001 | 0.905 |
| 144-155 | 5.185 | < .001 | 1.174 |
| 197-155 | 5.404 | < .001 | 1.224 |

| | | | |
|---|---|---|---|
| 109-155 | 4.516 | < .001 | 1.023 |
| 22-155 | 4.6 | < .001 | 1.042 |
| 70-155 | 5.776 | < .001 | 1.308 |

**Supplementary Figure 8: Scatters illustrating the brain-behavior correlations coloured by group** to supplement figure 4 – which shows heat map for all participants together. (A) basic auditory processing areas: left thalamus (ROIs 67, 113) to left (ROIs 144, 176) and right (ROIs 7, 185) Heschl's gyrus; (B) secondary auditory cortex areas: left (ROIs 144, 176) and right (ROIs 7, 185) Heschl's gyrus to left (ROIs 109, 197) and right (ROIs 22, 70) temporal areas; and frontal-to-temporal areas, moving in a dorsal-posterior direction, between left (ROIs 144, 197, 109) and right (ROIs 22, 70) temporal areas to (C) left frontal (ROIs 168, 48, 124, 157, 66, 179) and (D) right frontal (ROIs 60, 94, 59, 129, 110, 155) areas.

A.

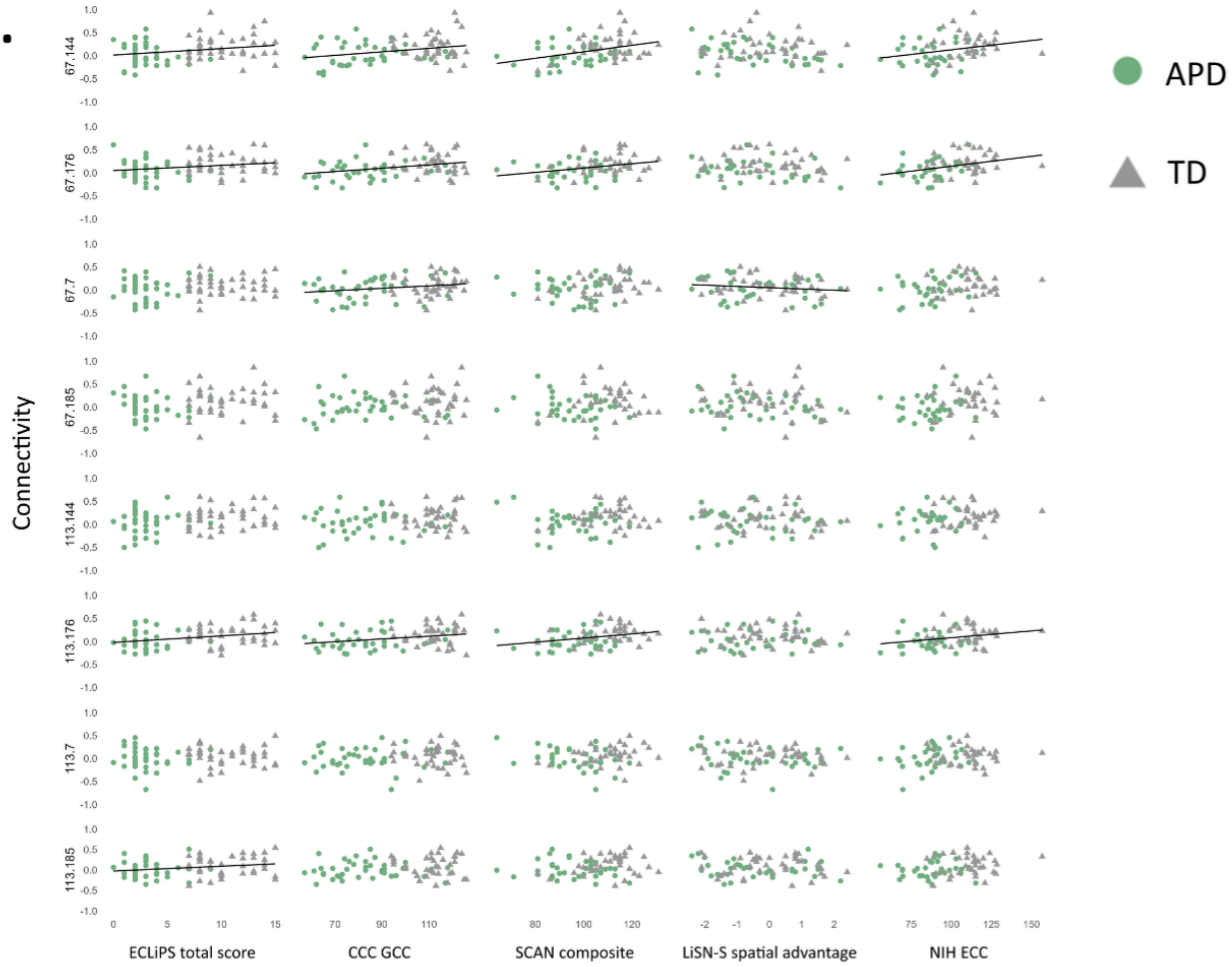

B.

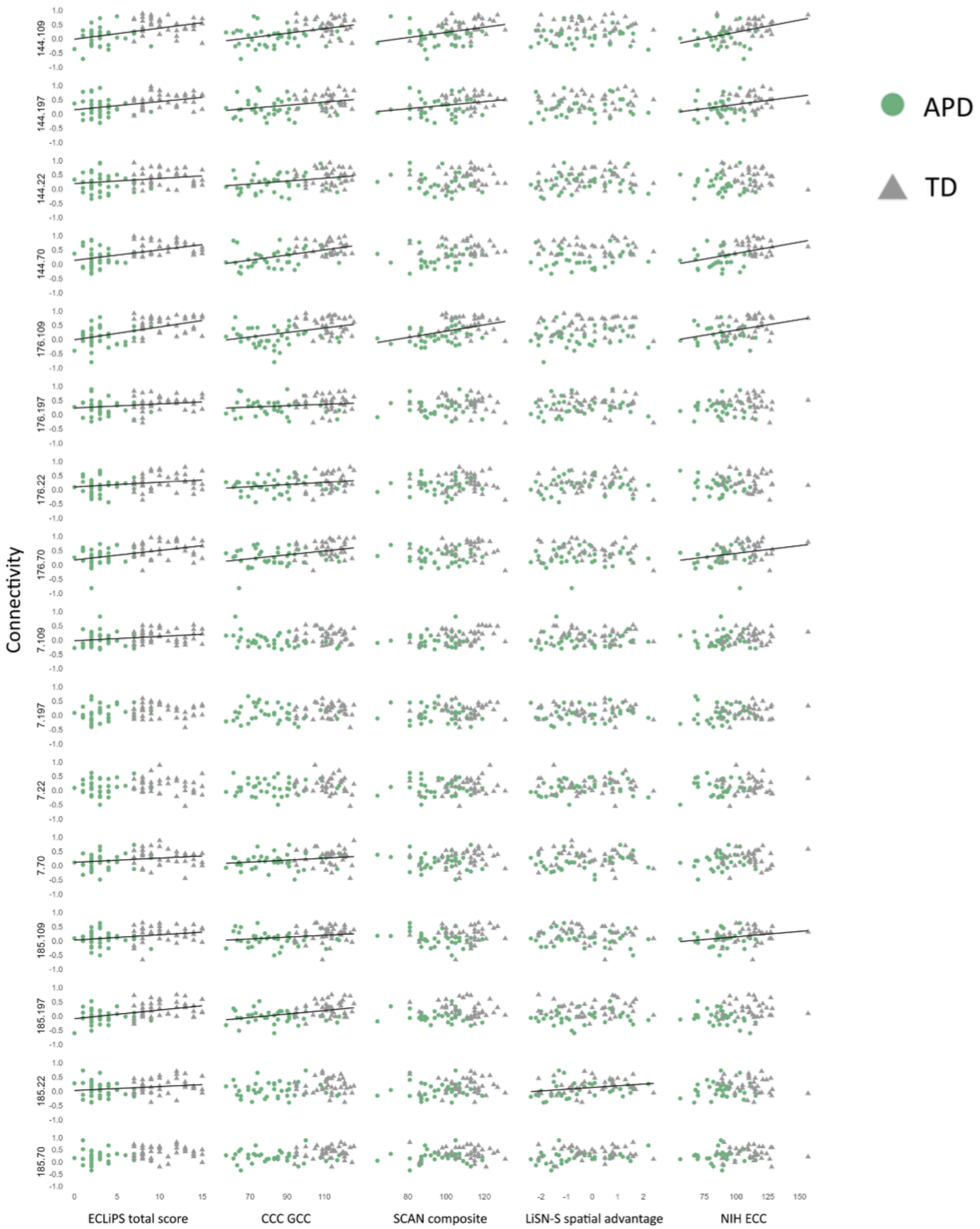

Connectivity
144.109
144.197
144.22
144.70
176.109
176.197
176.22
176.70
7.109
7.197
7.22
7.70
185.109
185.197
185.22
185.70
ECLiPS total score
CCC GCC
SCAN composite
LiSN-S spatial advantage
NIH ECC
APD
TD

Ci.

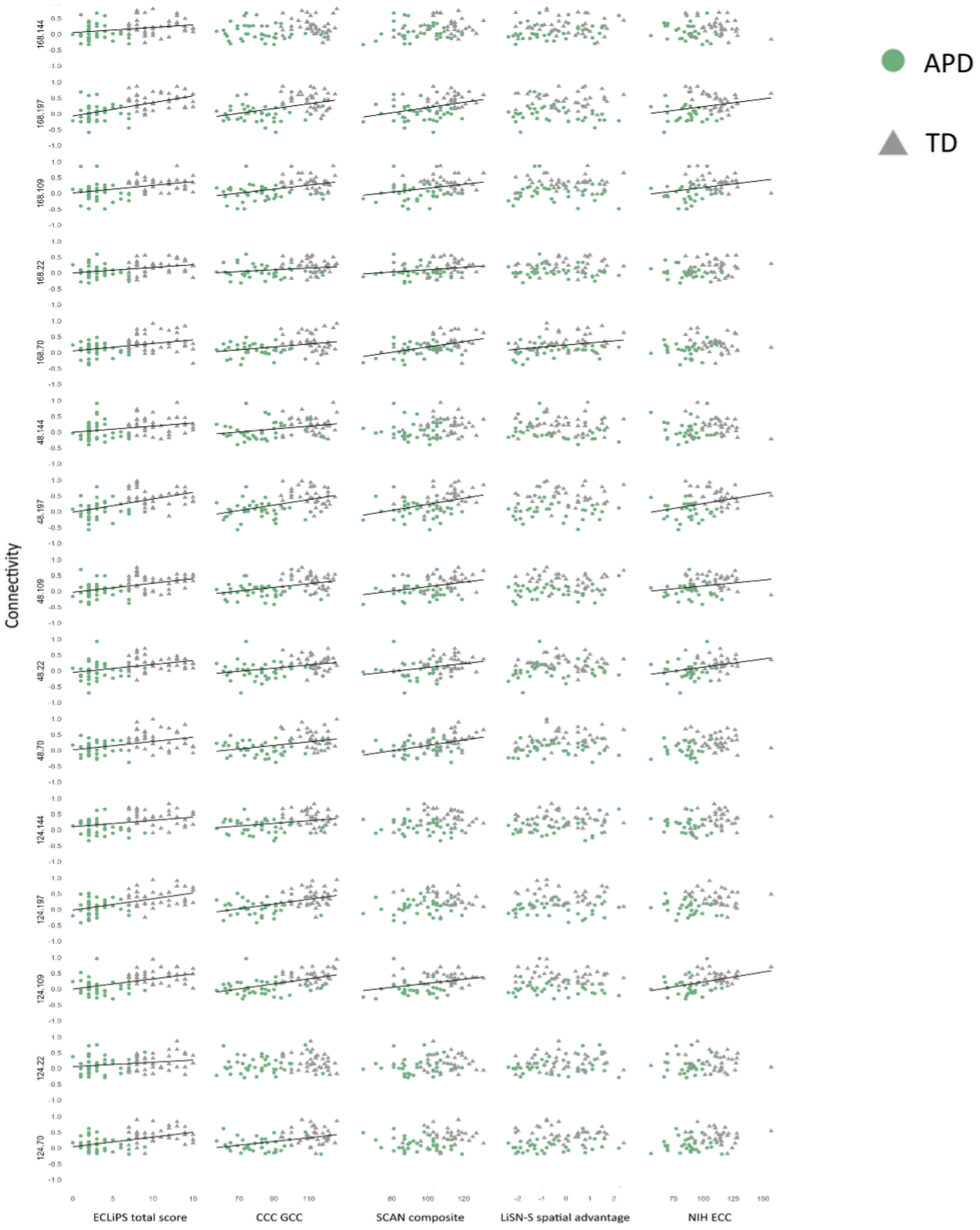

APD
TD
Connectivity
168.144
168.197
168.109
168.22
168.70
48.144
48.197
48.109
48.22
48.70
124.144
124.197
124.109
124.22
124.70
ECLiPS total score
CCC GCC
SCAN composite
LiSN-S spatial advantage
NIH ECC

Cii.

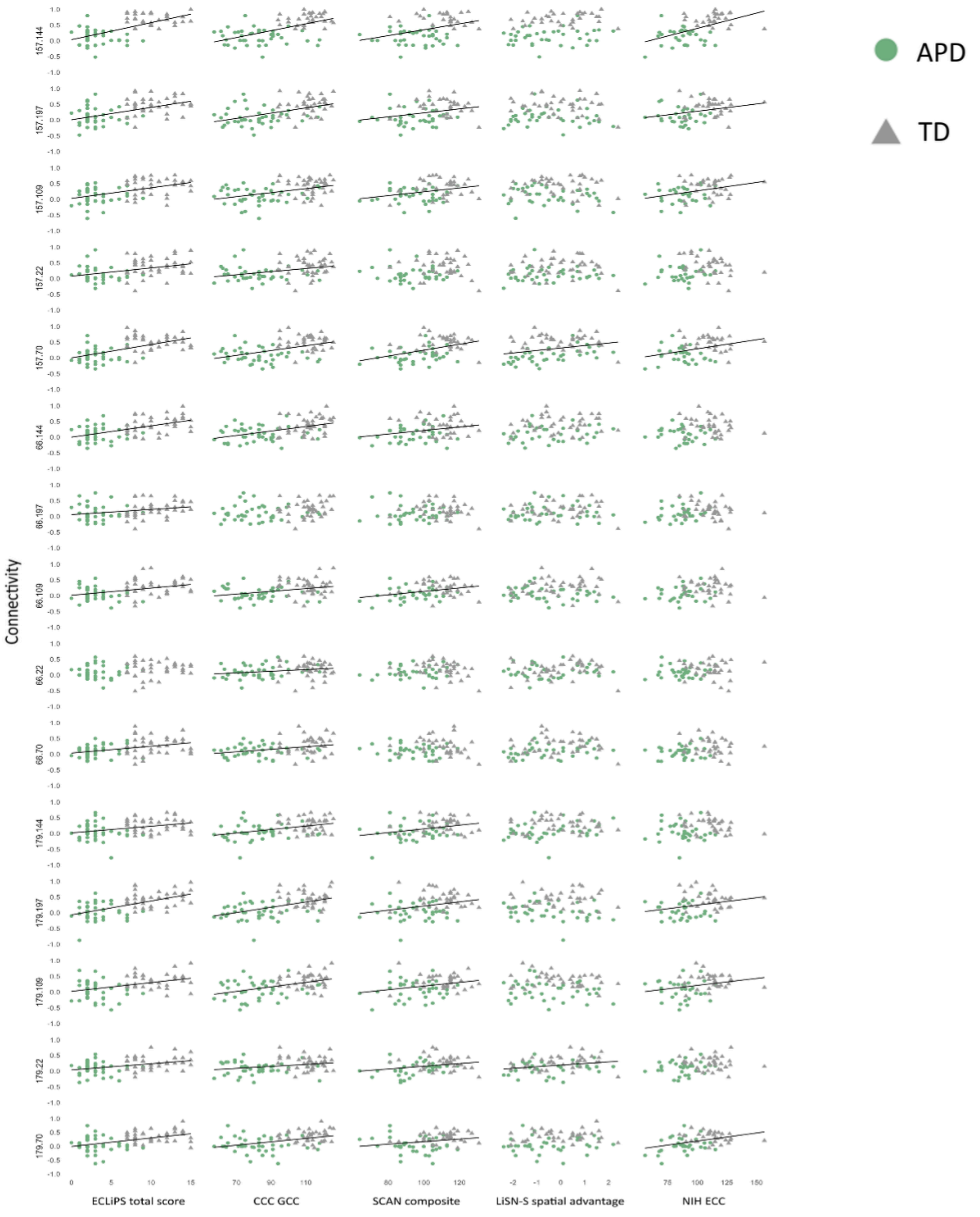

APD
TD
Connectivity
157.144
157.197
157.109
157.22
157.70
66.144
66.197
66.109
66.22
66.70
179.144
179.197
179.109
179.22
179.70
ECLiPS total score
CCC GCC
SCAN composite
LiSN-S spatial advantage
NIH ECC

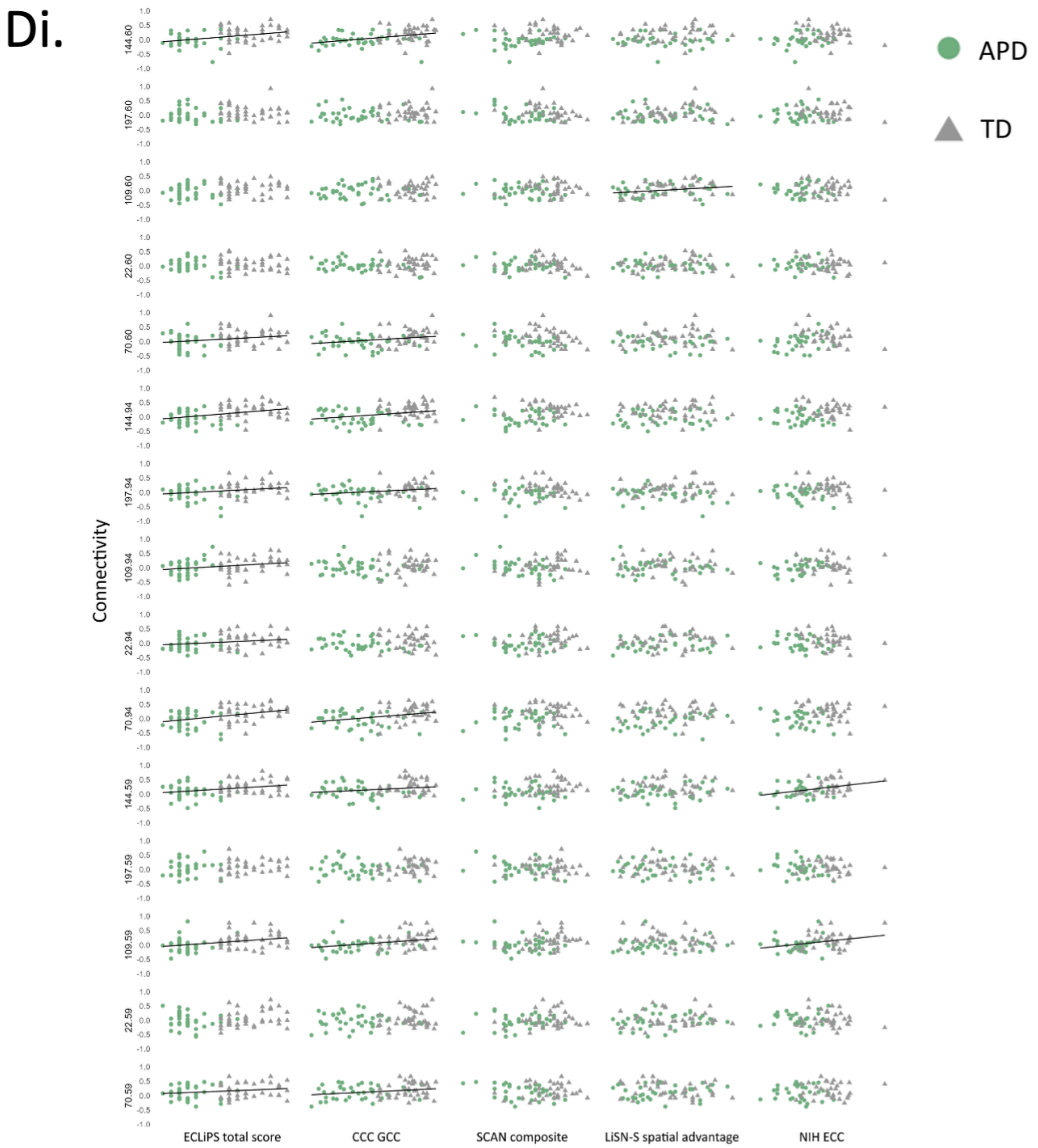

Di.
APD
TD
Connectivity
144.60
197.60
109.60
22.60
70.60
144.94
197.94
109.94
22.94
70.94
144.59
197.59
109.59
22.59
70.59
1.0
0.5
0.0
-0.5
-1.0
ECLiPS total score
CCC GCC
SCAN composite
LiSN-S spatial advantage
NIH ECC

Dii.

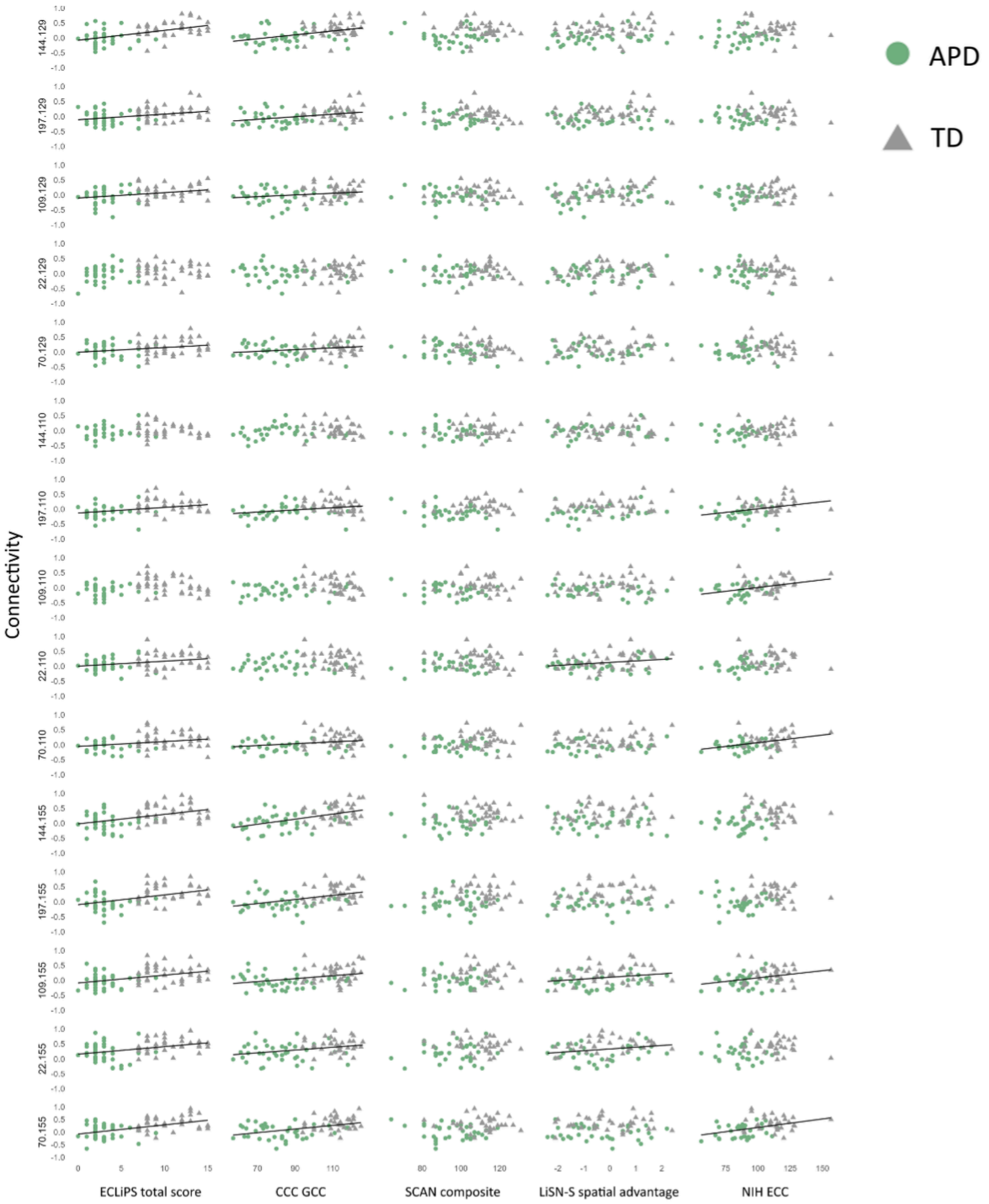
APD
TD
Connectivity
144.129
197.129
109.129
22.129
70.129
144.110
197.110
109.110
22.110
70.110
144.155
197.155
109.155
22.155
70.155
ECLiPS total score
CCC GCC
SCAN composite
LiSN-S spatial advantage
NIH ECC

**Supplementary Figure 9: Age related changes in speech and sound connectivity in all three groups.** ROI-ROI resting state connectivity for (A) Speech, (B) Sound and (C) Visual networks broken down into age groups for each participant group. Thicker, more saturated color lines represent stronger connections between cortical areas. TD children show significant increases and decreases in connectivity of the Speech network along with increases in connectivity in the Sound network until age 10/11. The children with LiD show big increases in connectivity in the Speech network in the younger age groups, along with smaller increases in the Sound network in the 8/9 and 10/11 y.o. groups. They then show a decrease in connectivity in both the Speech and Sound networks comparing the 10/11 y.o. to the 12/13 y.o.. The children with ADHD show increases in connectivity in both the Speech and Sound networks comparing the 8/9 y.o. to the 10/12 y.o. Only the TD children showed very minor increases in connectivity in the Visual network with age.

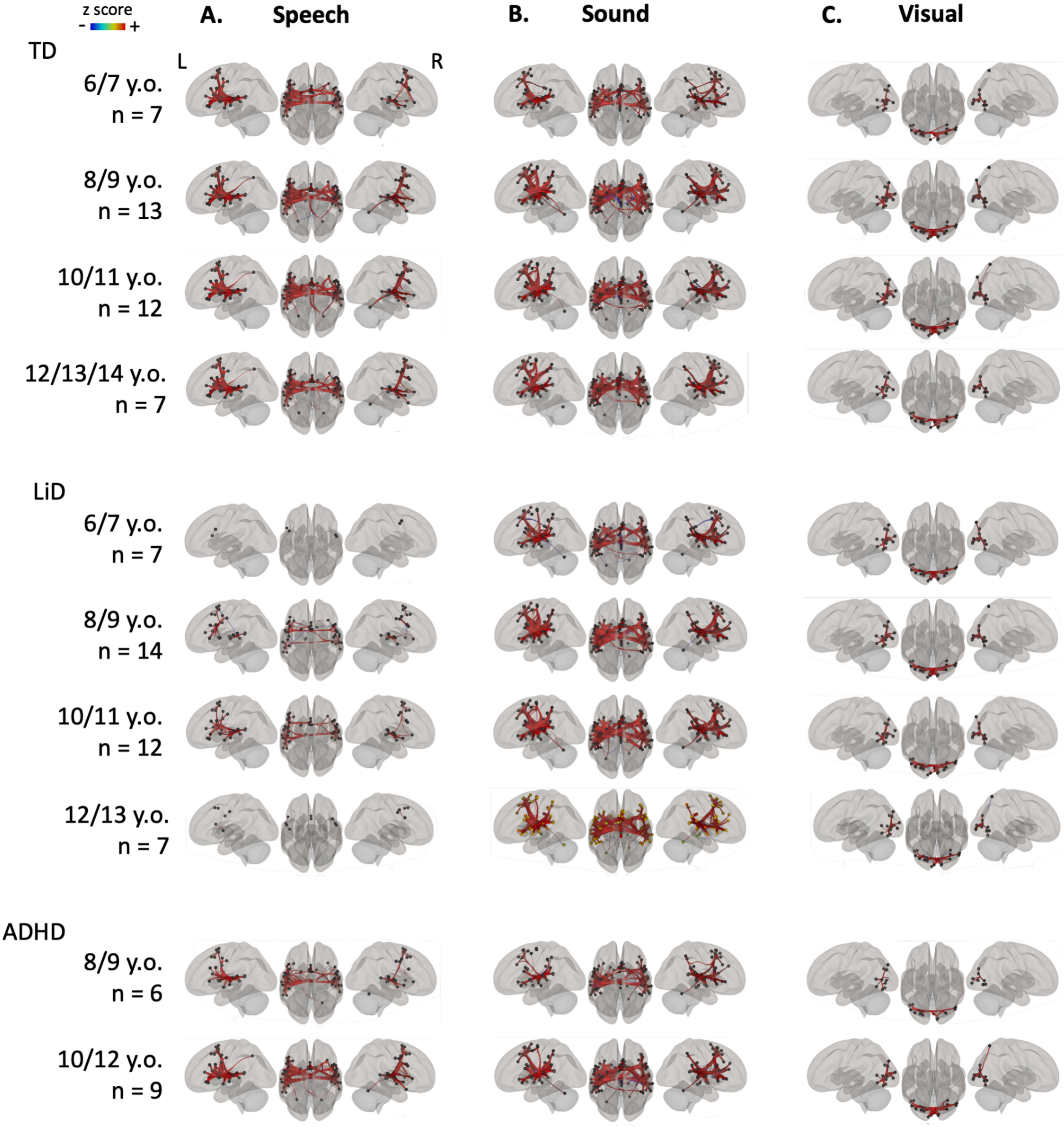
z score
-
+
A.
Speech
B.
Sound
C.
Visual
L
R
TD
6/7 y.o.
n = 7
8/9 y.o.
n = 13
10/11 y.o.
n = 12
12/13/14 y.o.
n = 7
LiD
6/7 y.o.
n = 7
8/9 y.o.
n = 14
10/11 y.o.
n = 12
12/13 y.o.
n = 7
ADHD
8/9 y.o.
n = 6
10/12 y.o.
n = 9

**Supplementary Table 5**: **Post-hoc GLMs compared age groups within each participant group for each of the three networks.** This is illustrated in Figure 5 and Supp. Fig. 7. ROI details are in Supp. Table 2.

| Network | Group | Age comparison | | ROI - ROI | df | t | p |
|---|---|---|---|---|---|---|---|
| Speech | TD | 6/7 | 8/9 | 12-23 | 18 | -3.87 | 0.0271 |
| | | | | 12-4 | 18 | -3.87 | 0.0271 |
| | | | | 49-1 | 18 | 4.09 | 0.0328 |
| | | | | 20-41 | 18 | 3.93 | 0.0475 |
| | | | 10/11 | 5-7 | 17 | 4.37 | 0.0199 |
| | | | 12/13/14 | 40-33 | 12 | 4.97 | 0.0145 |
| | | | | 40-18 | 12 | 4.61 | 0.0154 |
| | | 8/9 | 10/11 | 3-32 | 23 | 4.23 | 0.0154 |
| | | | | 3-7 | 23 | 3.85 | 0.0195 |
| | | | | 3-14 | 23 | 3.32 | 0.0464 |
| | | | | 3-20 | 23 | -3.21 | 0.0464 |
| | | | | 20-44 | 23 | -3.94 | 0.031 |
| | | | | 32-26 | 23 | 3.66 | 0.0316 |
| | | | | 31-23 | 23 | -3.88 | 0.0361 |
| | | | 12/13/14 | n.s. | | | |
| | | 10/11 | 12/13/14 | n.s. | | | |
| | LiD | 6/7 | 8/9 | 44-28 | 20 | 4.75 | 0.0059 |
| | | | | 5-48 | 20 | 3.94 | 0.0388 |
| | | | 10/11 | 27-8 | 16 | 4.46 | 0.0189 |
| | | | | 27-34 | 16 | 3.88 | 0.0321 |
| | | | | 26-44 | 16 | 4.37 | 0.0229 |

| | | | | | | | |
|---|---|---|---|---|---|---|---|
| | | | 12/13 | 3-24 | 12 | 4.47 | 0.0368 |
| | | 8/9 | 10/11 | 8-14 | 24 | 3.8 | 0.021 |
| | | | | 8-13 | 24 | 3.39 | 0.0382 |
| | | | | 8-27 | 24 | 3.88 | 0.021 |
| | | | | 27-34 | 24 | 3.67 | 0.029 |
| | | | | 35-24 | 24 | 3.85 | 0.0372 |
| | | | | 44-31 | 24 | 3.8 | 0.0361 |
| | | | | 44-26 | 24 | 3.58 | 0.0361 |
| | | | 12/13 | 13-15 | 20 | 5.45 | 0.0012 |
| | | | | 21-28 | 20 | -5.41 | 0.0013 |
| | | | | 42-4 | 20 | -3.88 | 0.0223 |
| | | | | 42-24 | 20 | 4.46 | 0.0015 |
| | | | | 11-7 | 20 | -4.28 | 0.0175 |
| | | | | 35-14 | 20 | 4.36 | 0.0145 |
| | | 10/11 | 12/13 | 19-39 | 16 | -4.51 | 0.0395 |
| | ADHD | 8/9 | 10/11 | ns | | | |
| | | | 12/13 | ns | | | |
| | | 10/11 | 12/13 | 19-47 | 7 | 5.76 | 0.0332 |
| | | | | 16-38 | 7 | -4.96 | 0.0476 |
| | | | | 16-48 | 7 | 4.79 | 0.0476 |
| Sound | TD | 6/7 | 8/9 | 54-3 | 18 | -4.22 | 0.0348 |
| | | | | 67-12 | 18 | 4.1 | 0.0392 |
| | | | | 67-65 | 18 | -3.8 | 0.0392 |
| | | | | 67-3 | 18 | -3.68 | 0.0392 |
| | | | | 23-37 | 18 | -4.64 | 0.0138 |

| | | | | | |
|---|---|---|---|---|---|
| | 10/11 | 3-67 | 17 | -4.76 | 0.0107 |
| | | 3-14 | 17 | -4.5 | 0.0107 |
| | | 3-9 | 17 | -4.02 | 0.0201 |
| | | 2-33 | 17 | 5.01 | 0.0073 |
| | | 2-44 | 17 | 4.24 | 0.0186 |
| | | 2-8 | 17 | 3.94 | 0.024 |
| | | 16-6 | 17 | 4.11 | 0.0494 |
| | | 33-19 | 17 | 5.08 | 0.0036 |
| | | 33-2 | 17 | 5.01 | 0.0036 |
| | | 33-8 | 17 | 3.87 | 0.0276 |
| | | 59-46 | 17 | 4.49 | 0.022 |
| | | 8-6 | 17 | 4.28 | 0.0276 |
| | | 8-59 | 17 | 3.54 | 0.0427 |
| | | 8-19 | 17 | 3.38 | 0.0488 |
| | | 6-16 | 17 | 4.11 | 0.0247 |
| | | 65-67 | 17 | -4.5 | 0.0215 |
| | | 65-14 | 17 | -3.98 | 0.0328 |
| | 12/13/14 | 66-54 | 12 | 5.92 | 0.0048 |
| 8/9 | 10/11 | 8-2 | 23 | 4.7 | 0.0067 |
| | | 8-26 | 23 | 3.95 | 0.0189 |
| | | 8-6 | 23 | 3.77 | 0.0189 |
| | | 8-4 | 23 | 3.72 | 0.0189 |
| | | 16-2 | 23 | 3.99 | 0.0393 |
| | | 2-38 | 23 | 4.84 | 0.0034 |
| | | 2-33 | 23 | 3.78 | 0.0165 |

| | | | | | | |
|---|---|---|---|---|---|---|
| | | | 2-44 | 23 | 3.6 | 0.0207 |
| | | | 2-19 | 23 | 3.38 | 0.0294 |
| | | | 6-16 | 23 | 3.46 | 0.0447 |
| | | | 6-20 | 23 | 3.42 | 0.0447 |
| | | | 6-11 | 23 | 3.35 | 0.0447 |
| | | | 6-56 | 23 | 3.16 | 0.049 |
| | | | 6-22 | 23 | 3.1 | 0.049 |
| | | 12/13/14 | 42-19 | 18 | 4.07 | 0.0494 |
| | 10/11 | 13/13/14 | 53-45 | 17 | 4.74 | 0.0128 |
| LiD | 6/7 | 8/9 | ns | | | |
| | | 10/11 | ns | | | |
| | | 12/13 | 18-57 | 12 | -5.97 | 0.0044 |
| | | | 40-4 | 12 | -5.42 | 0.0105 |
| | | | 40-14 | 12 | -4 | 0.0416 |
| | | | 40-39 | 12 | -3.98 | 0.0416 |
| | | | 58-69 | 12 | 4.62 | 0.0399 |
| | | | 5-1 | 12 | -4.38 | 0.0463 |
| | | | 5-50 | 12 | -4.14 | 0.0463 |
| | 8/9 | 10/11 | 47-63 | 24 | 4.72 | 0.0057 |
| | | | 47-49 | 24 | 4.26 | 0.0093 |
| | | | 67-64 | 24 | 4.18 | 0.0229 |
| | | | 67-11 | 24 | 3.79 | 0.0302 |
| | | 12/13 | ns | | | |
| | 10/11 | 12/13 | ns | | | |
| ADHD | 8/9 | 10/11 | 65-67 | 10 | -5.13 | 0.0171 |

| | | | | | | | |
|---|---|---|---|---|---|---|---|
| | | | | 65-7 | 10 | -5.05 | 0.0171 |
| | | | 12/13 | 69-11 | 5 | 8.29 | 0.0284 |
| | | 10/11 | 12/13 | 13-46 | 7 | 6.5 | 0.0226 |
| Visual | TD | 6/7 | 8/9 | 1-11 | 18 | 4.12 | 0.0134 |
| | | | 10/11 | ns | | | |
| | | | 12/13/14 | ns | | | |
| | | 8/9 | 10/11 | 4-21 | 23 | -3.59 | 0.0321 |
| | | | 12/13/14 | ns | | | |
| | | 10/11 | 12/13/14 | 17-20 | 17 | -3.95 | 0.0218 |
| | LiD | 6/7 | 8/9 | ns | | | |
| | | | 10/11 | ns | | | |
| | | | 12/13 | ns | | | |
| | | 8/9 | 10/11 | ns | | | |
| | | | 12/13 | ns | | | |
| | | 10/11 | 12/13 | ns | | | |
| | ADHD | 8/9 | 10/11 | ns | | | |
| | | | 12/13 | ns | | | |
| | | 10/11 | 12/13 | ns | | | |
